# Long-term morphometric similarity gradients relate to cortical hierarchy and psychiatric symptoms in schizophrenia

**DOI:** 10.64898/2026.02.25.26347075

**Authors:** Natalia García-San-Martín, Richard AI Bethlehem, Isaac Sebenius, Leonardo Cardoso Saraiva, Patricia Segura, Claudio Alemán-Morillo, Chloé Gomez, Pablo Salguero-Quirós, Alessia Pasquini, Marcella Montagnese, Golia Shafiei, Miguel Ruiz-Veguilla, Rosa Ayesa-Arriola, Javier Vázquez-Bourgon, Bratislav Misic, Carolina Cappi, John Suckling, Benedicto Crespo-Facorro, Rafael Romero-García

**Affiliations:** Department of Medical Physiology and Biophysics, Faculty of Medicine, University of Seville; Seville, Spain; Department of Psychology, University of Cambridge; Cambridge, UK; Department of Psychiatry, University of Cambridge; Cambridge, UK; Harvard Medical School; Boston, Massachusetts, MA, USA; Department of Psychiatry, Yale University School of Medicine; New Haven, Connecticut, CT, USA; Department of Psychiatry, Perelman School of Medicine, University of Pennsylvania; Philadelphia, PA, USA; Penn Lifespan Informatics and Neuroimaging Center (PennLINC), Perelman School of Medicine, University of Pennsylvania; Philadelphia, PA, United States; Lifespan Brain Institute (LiBI), The Children’s Hospital of Philadelphia and Penn Medicine; Philadelphia, PA, USA; Biomedical Research Center in Mental Health Network (CIBERSAM), Health Institute Carlos III; Madrid, Spain; Mental Health Service, Virgen del Rocío University Hospital; Seville, Spain; Instituto de Biomedicina de Sevilla (IBiS) HUVR/CSIC, University of Seville; Seville, Spain. Department of Psychiatry, Marqués de Valdecilla University Hospital, IDIVAL, School of; Medicine, University of Cantabria; Santander, Spain; Montreal Neurological Institute, McGill University; Montreal, QC, Canada; Department of Psychiatry, Robert Wood Johnson Medical School; New Brunswick, NJ, USA; Peterborough NHS Foundation Trust; Peterborough, UK

## Abstract

Schizophrenia spectrum disorders (SSD) are characterized by altered brain structure, reflecting widespread dysconnectivity across brain-specific networks. However, the role of hierarchical organization on cortical morphometric networks in shaping clinical outcomes over the course of the disease remains unclear. Connectome-derived gradients have increasingly been used to investigate spatial transitions in brain organization. Here, we computed cortical and subcortical Morphometric INverse Divergence (MIND) similarity networks from 1293 structural MRI data of 193 healthy controls (HC) and 350 individuals with SSD followed for up to 20 years. MIND features were calculated for each subject-specific network by computing regional averages and performing gradient decomposition. We found that MIND was longitudinally associated with treatment duration and medication in SSD. These associations were co-localized with hierarchical axes of cortical organization and schizophrenia epicenters. Moreover, psychiatric symptoms were associated with these alterations in structural similarity, which were also related to treatment duration. Collectively, these findings advance our understanding of how brain organization, treatment duration, and medication shape clinical symptoms throughout the course of SSD.

## Introduction

The initial phases of schizophrenia spectrum disorders (SSD) are characterized by brain structural alterations [1], which have been shown to predict long-term clinical trajectories following first episode psychosis [2, 3], highlighting their relevance for understanding disease onset and progression. Accordingly, longitudinal studies have also revealed that brain tissue loss persists during the years following illness onset, including gray matter reductions [4] and progressive cortical thinning [5] over time. These changes likely reflect aberrant neurodevelopmental processes that may occur at various stages of life [6, 7]. Furthermore, the psychotic episode itself—along with the associated antipsychotic treatment—may also contribute to additional alterations in brain morphology [8]. Disentangling the impact of disease-related mechanisms from medication effects on brain structure remains a major challenge [9, 10]. Thus, the temporal course of these alterations across the psychosis spectrum remains inconclusive [11], likely influenced by the clinical heterogeneity of symptoms and the effects of antipsychotic drugs [9].

The morphological brain alterations related to schizophrenia (SCZ) are constrained by the underlying structural connectome, which comprises a network of functionally specialized regions interconnected by axonal pathways [12, 13]. In this context, Morphometric INverse Divergence (MIND) is a recently developed method that estimates within-subject similarity between brain regions by quantifying the divergence between multivariate distributions of multiple MRI-derived features [14, 15]. It provides a useful metric of structural similarity—a proxy for connectivity—derived from cortical vertex-level surface reconstructions, thereby overcoming the practical constraints associated with acquiring multimodal MRI data [16]. Although cortical MIND studies have identified patterns of brain reorganization in neurodevelopmental [17] and neurodegenerative [18] conditions, the absence of generalized cortical and subcortical connectome-based biomarkers for assessing illness progression further complicates the prediction of long-term symptom severity [19]. Moreover, the wide heterogeneity in symptom profiles and clinical trajectories makes it even more challenging to establish consistent patterns of structural change across individuals. Characterizing the impact of atypical development on network architecture is thus essential for understanding clinical variability in neuropsychiatric disorders [20].

Recent advances in human connectomics have revealed multiple large-scale brain networks characterized by distinct functional processing profiles [21], including those involved in basic sensory and motor (primary) functions, attentional or cognitive control processes [22], and higher-order networks at the top of the cortical hierarchy, such as the default mode network (DMN) [23]. Exploring the differentiation of these connectivity patterns enables a more detailed characterization of hierarchical brain organization [24]. This differentiation is commonly achieved using gradient decomposition, which reduces high-dimensional connectivity features into low-dimensional representations. Connectome-derived gradients are often revealed across distinct processing pathways or streams. For example, within the visual system, information transitions from simple visual features encoded in the primary visual cortex to increasingly complex representations encoded in temporal regions, ultimately supporting high-level semantic processing [25]. Thus, gradients provide a systematic mapping between spatial organization and the hierarchical continuum of information processing. For instance, in both humans and the macaque monkey, the decomposition of functional connectivity data has revealed a functional gradient that transitions from regions supporting primary sensory and motor functions to transmodal regions, which in humans overlap with components of the DMN. A recent study has extended this framework by establishing a continuous normative reference to explore how these functional gradients evolve across the lifespan, revealing that gradient architecture is initially anchored by primary sensory systems, differentiates along association and control axes during adolescence, and progressively dedifferentiates with ageing [26]. Moreover, the decomposition of structural connectivity in neuropsychiatric disorders has revealed associations with neurotransmitters and cognition in bipolar disorder using MIND [27], as well as with clinical symptoms in schizophrenia using a structural covariance approach alternative to MIND [28]. However, the characterization of structural connectivity profiles in relation to the progression of schizophrenia remains under debate, and structural gradients of cortical organization are still largely unexplored.

The spatial distribution of psychosis-related alterations is not uniform across the cortex but instead preferentially affects regions involved in higher-order functions, including those shaped by evolutionary expansion [29] as well as functional [30] and association [31] processes. In particular, the cortical pattern of schizophrenia-related dysconnectivity has been shown to overlap with evolutionary modifications of human brain connectivity [29]. This altered connectivity supporting higher-order brain functions may also increase susceptibility to brain dysfunction, suggesting that human brain evolution and hierarchical organization may contribute to region-specific vulnerability in the disorder. Consistent with this view, studies mapping neurobiological features [32] and disease epicenters [33] in schizophrenia have revealed that certain regions exhibit greater vulnerability to the disease. Furthermore, these regional alterations have also been associated with symptom severity and antipsychotic treatment [4, 9, 30]. Disentangling region-specific disease progression from brain organizational processes and medication effects is therefore essential for clarifying the role of brain connectivity in schizophrenia symptomatology.

To our knowledge, no previous study has examined how cortical gradients evolve longitudinally in SSD or their relationship with hierarchical brain organization and psychiatric symptomatology over time. Here, we used cortical and subcortical structural networks to assess how brain organization, captured by MIND degree and gradients, is associated with diagnosis, treatment duration, medication, and symptomatology in SSD, and how these alterations co-localize with hierarchical axes of cortical organization and SCZ epicenters.

## Subjects and Methods

### Subjects

Magnetic Resonance Imaging (MRI) data from individuals meeting clinical criteria for a first-episode non-affective psychosis, consistent with a schizophrenia spectrum disorder (SSD, *n* = 365, *n*_female_ = 145, *age* = 29.69 ± 8.76), along with healthy controls (HC, *n* = 196, *n*_female_ = 75, *age* = 29.04 ± 7.61), were obtained from *Programa de Atención a las Fases Iniciales de Psicosis* (PAFIP) [34]. Individuals with SSD were either antipsychotic-naïve or had received minimal antipsychotic treatment (< 6 weeks of antipsychotics). All diagnoses were made by an experienced psychiatrist using the Structured Clinical Interview for DSM-IV (SCID-I), confirming the presence of schizophrenia (SCZ) or other SSD after six months of the first visit (see Supplementary *Subjects and Methods* for additional inclusion and exclusion criteria). Participants provided written informed consent and underwent follow-up assessments at 1, 3, 5, 10, 15, and 20 years.

This program has been approved by the CEIC-Cantabria Ethics Committee for Clinical Research (NCT0235832) and the University of Seville Ethics Committee (SICEIA 2024-2534) in accordance with international standards.

### MRI acquisition, parcellation, and volume extraction

T_1_-weighted images acquired from two 1.5T and 3T MRI scanners were processed using the FreeSurfer 7.4.1 longitudinal stream [35], applying the recon-all pipeline. All images underwent a quality control (QC) protocol. Images were visually inspected by trained raters, resulting in a total of 1293 good quality images. As all remaining images had a cortical surface reconstruction quality metric—based on topological defects—within the mean ± 5 standard deviations (Euler number > -135), no further participants were excluded.

Cortical brain parcellation was performed using a symmetric, equally-sized (500 mm^2^) sub-parcellation of the Desikan-Killiany (DK) atlas [36], resulting in 318 cortical regions-of-interest. 14 subcortical structures were also considered for further analyses (bilateral accumbens, amygdala, caudate, hippocampus, pallidum, putamen, and thalamus). See Supplementary *Subjects and Methods* for detailed MRI acquisition parameters and the follow-up periods after QC. See Supplementary Figs. 1 and 2 for the flow diagram of study participants and the alluvial diagram of attrition, respectively.

**Fig. 1.**
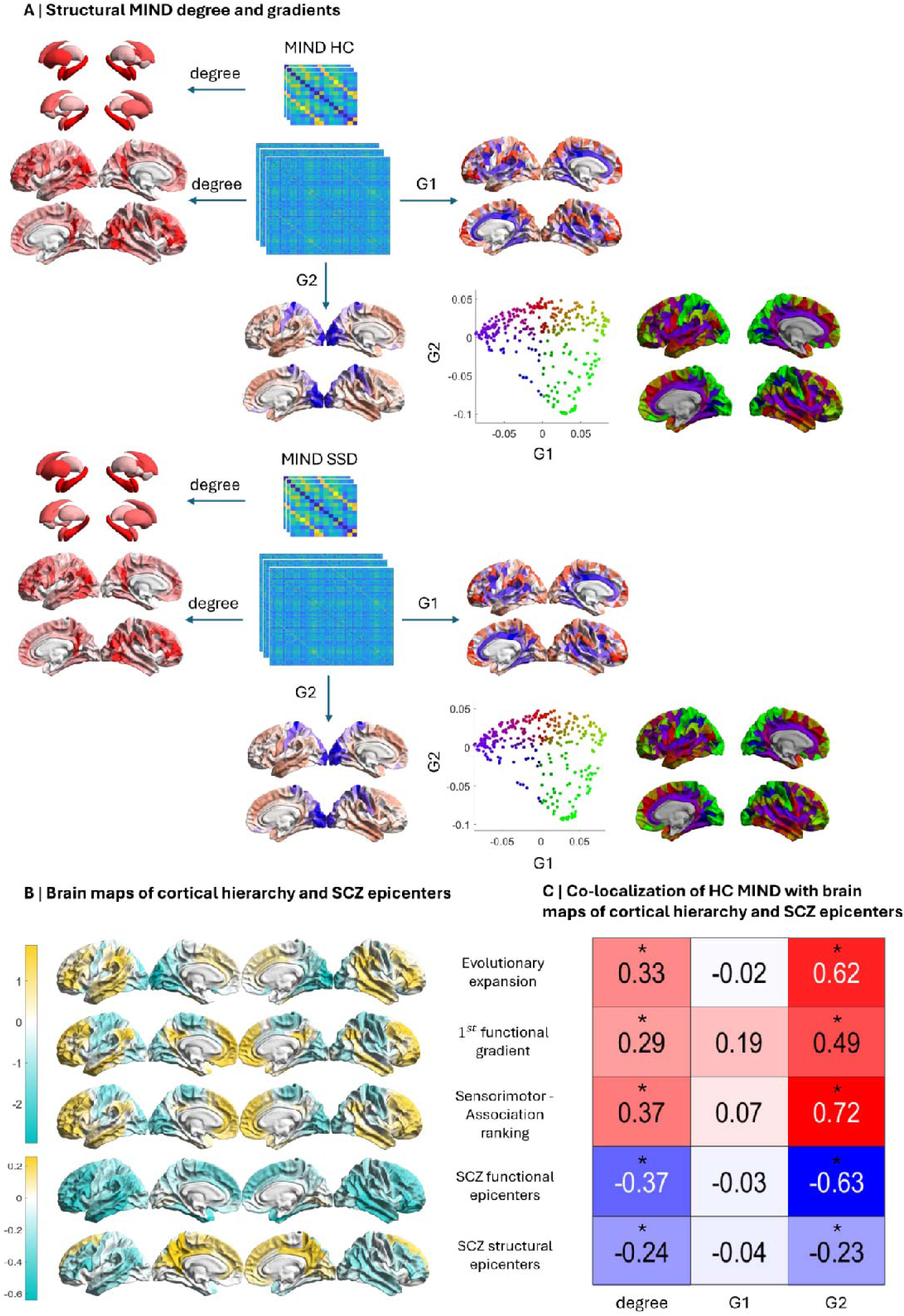
Degrees and gradients of structural connectivity derived from MIND networks. (**A**) Schematic analysis pipeline. Cortical and subcortical degree, along with cortical gradients (G1 and G2), were computed from MIND networks of each individual, separately for HC and SSD groups. Scatter plots show the first two principal gradients of structural connectivity—obtained using gradient decomposition—derived from the group-averaged MIND matrices of each individual. Gradients are also plotted on the cortical surface, with colour representing the position of each region along each gradient. (**B**) Brain maps of evolutionary, functional, and S-A ranking features were derived from *neuromaps* [40] (top); the spatial maps of functional and structural epicenters in SCZ were obtained from an ENIGMA study (bottom) [33]. **(C**) Regional co-localization of cortical MIND features (degree, G1, and G2) in HC with brain maps of cortical hierarchy and SCZ epicenters. Asterisks (*) indicate significant correlations (Benjamini-Hochberg false discovery rate (FDR)-corrected across features; *P*_spin_<□0.05). G1, first gradient; G2, second gradient; HC, healthy control; MIND, morphometric inverse divergence; SCZ, schizophrenia; SSD, schizophrenia spectrum disorders.

**Fig. 2.**
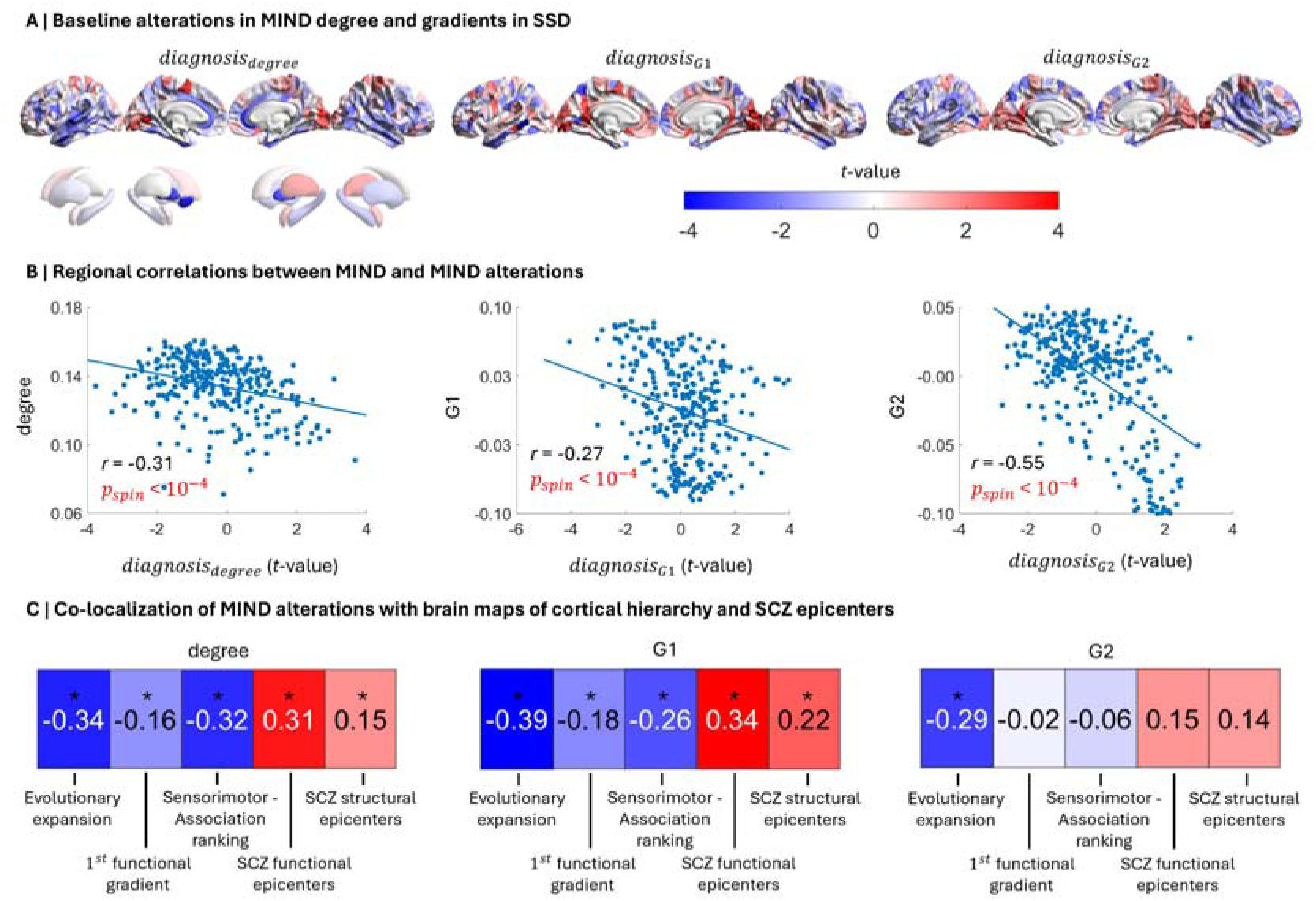
Baseline alterations in MIND degree and gradients in SSD associated with hierarchical organization and SCZ epicenters. (**A**) Regional associations (*t*-values) representing the contribution of the effect of diagnosis on brain similarity at baseline assessed by multiple regression models (*diagnosis*; from left to right: degree, G1, and G2; FDR-corrected, *P*□<□0.05). (**B**) Regional correlations (two-sided test) between the effect of diagnosis on brain similarity (*diagnosis*) and brain similarity in HC. (**C**) Regional co-localization (two-sided test) of the effect of diagnosis on brain similarity (*diagnosis*) with brain maps of cortical hierarchy and SCZ epicenters, while accounting for regional brain similarity. Asterisks (*) indicate significant correlations (FDR-corrected across features; *P*_spin_<□0.05). G1, first gradient; G2, second gradient; HC, healthy control; MIND, morphometric inverse divergence; SCZ, schizophrenia; SSD, schizophrenia spectrum disorders.

### Cortical and subcortical MIND estimation

Morphometric INverse Divergence (MIND) networks were used to capture structural similarity between cortical and subcortical areas, separately. For each subject, a MIND network was constructed by computing between pairs of regions the Kullback-Leibler divergence of multiple MRI features (Supplementary *Subjects and Methods* for details) [14, 37]. This resulted in two edge-level matrices of size 318x318 and 14x14 bounded between 0 and 1, representing the structural connectivity between each pair of cortical and subcortical regions, respectively (Fig. 1A). To correct for site-related batch effects while preserving the effects of age, sex, diagnosis, and the diagnosis-by-sex interaction, we used ComBatLS [38], a recent extension of the ComBat method that preserves biological variance while reducing biases. Individual-level MIND degrees were computed by averaging their corresponding MIND edges, resulting in an average within-subject similarity degree for each region.

### Cortical MIND gradients

The first two structural connectivity gradients G1 and G2 were derived from cortical MIND networks (318x318) of each individual using gradient decomposition [39] (see Supplementary *Subjects and Methods* for details). Gradients from subcortical MIND networks (14x14) were not derived due to an insufficient number of variables to perform decomposition. Individual-level gradient ranges were computed for both G1 and G2 by computing the difference between the maximum and minimum values of their corresponding MIND gradients.

### Baseline and longitudinal MIND associations in SSD

To evaluate the impact of SSD diagnosis on brain similarity (degree and gradients) at baseline, multiple regression analyses were performed for each region, as shown in Eq. (1).

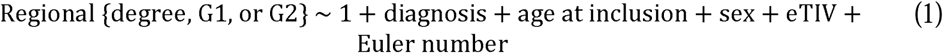

where ‘*eTIV*’ is the estimated total intracranial volume and ‘*Euler number*’ is the FreeSurfer’s Euler index, a proxy of motion-induced image artefacts.

The longitudinal effects of diagnosis, time, and medication on brain similarity were assessed using linear mixed modeling (LMM) as shown in Eq. (2).

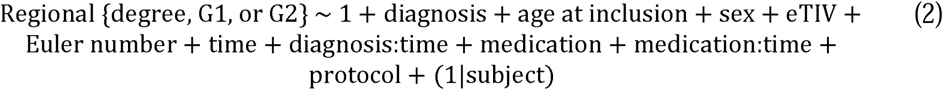

where ‘*time*’ represents the period since the enrolment for HC, or since treatment initiation for SSD; ‘*diagnosis:time*’ represents treatment duration for SSD individuals; ‘*medication*’ corresponds to antipsychotic dose expressed in chlorpromazine equivalents; ‘*protocol*’ is a binary variable that represents a change in the flip angle during MRI acquisition; and “*(1|subject)*” specifies a random intercept to account for subject-specific variability in overall offsets. For HC, ‘*medication*’ was set to zero. See Supplementary *Subjects and Methods* for definitions of the remaining variables. All non-binary variables were *z*-scored, and Benjamini-Hochberg false discovery rate (FDR) correction was applied across brain regions to account for multiple comparisons (*P* < 0.05). In addition to the region-level analyses, Eq. (1 and 2) were also applied to assess the effects of diagnosis, time, and medication on the gradient ranges of G1 and G2.

### Spatial co-localization of MIND with brain maps of cortical hierarchy and SCZ epicenters

Cortical brain maps of evolutionary expansion, the functional gradient, and the sensorimotor-association (S-A) ranking—collectively referred to here as hierarchical maps—were derived from *neuromaps* toolbox [40] and parcellated according to the same symmetric, equally-sized sub-parcellation of the DK atlas (318x318; Fig. 1B). Additionally, brain maps of functional and structural epicenters in SCZ were obtained from an ENIGMA study [33], and each value in the DK atlas was assigned to its correspondent region in the equally-sized DK sub-parcellation. See Supplementary *Subjects and Methods* for details on brain map definitions.

To investigate potential relationships between these brain maps and brain similarity, spatial co-localization was quantified by correlating (Pearson’s *r*) each MIND feature (cortical degree and gradients) with each brain map using a spin test (*n* = 10,000) that preserves spatial autocorrelation (see Supplementary *Subjects and Methods* for details). Similarly, we assessed the co-localization of the brain maps with the *t*-values derived from the baseline and longitudinal MIND models after accounting for their corresponding regional degree, G1, or G2, using the spin test and performing FDR correction across features.

### Baseline and longitudinal associations between MIND and symptomatology

MIND degree and gradients were additionally associated with psychiatric symptomatology in patients. General psychopathology was assessed using the expanded version of 24 items of the Brief Psychiatric Rating Scale (BPRS) [41]. Given that symptom scores are discrete count variables, baseline associations between brain similarity and BPRS scores were assessed using a generalized linear model (GLM) with a Poisson distribution (Eq. 3).

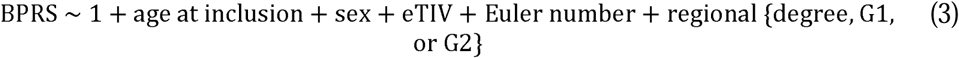

The longitudinal effect of brain similarity, treatment duration, and medication on BPRS scores was assessed using a generalized LMM (Eq. 4).

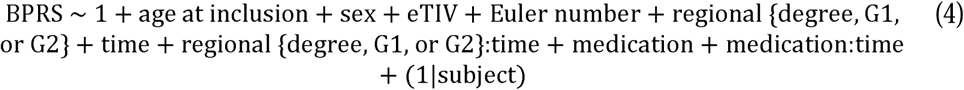

where ‘*regional{degree, G1, or G2}:time*’ corresponds to the interaction between the corresponding MIND feature and time. The ‘*protocol*’ effect was not included because all individuals with BPRS scores followed the same MRI protocol. All non-binary variables were *z*-scored, and FDR correction was applied across regions.

## Results

### Regional MIND is co-localized with brain maps of cortical hierarchy and SCZ epicenters

Cortical and subcortical MIND networks were computed for each participant, representing the within-subject similarity based on multiple MRI features (Fig. 1A). MIND degree exhibited higher similarity in prefrontal, inferior parietal, and supramarginal regions in both SSD and HC groups. Structural gradients were derived from cortical MIND networks, yielding representations of the brain’s spatial organization along its hierarchical spectrum. The first two gradients (G1 and G2) explained 16.4% and 15.1% of variance in MIND, respectively. While G1 showed a pattern anchored at one end by the cingulate and superior temporal cortices and at the other end by prefrontal, superior parietal, and occipital regions, G2 notably captured the sensorimotor-to-association (S-A) transition (see *Source Data* for degree, G1, and G2 of HC and SSD individuals at baseline). Degree and G2 showed a positive correlation with evolutionary expansion (Fig. 1C; Pearson’s *r* = 0.33 and *r* = 0.62, respectively; *P*_spin_<10^-4^), the functional gradient (*r* = 0.29, *P*_spin_=5·10^-4^; *r* = 0.49, *P*_spin_<10^-4^), and the S-A ranking (*r* = 0.37; *r* = 0.72; *P*_spin_<10^-4^), indicating that regions with higher structural similarity in association areas exhibit greater functional connectivity and evolutionary expansion. On the other hand, G1 did not show any significant co-localization.

Additionally, we found a negative correlation for both degree and G2 with the functional (*r* = -0.37 and *r* = -0.63, respectively; *P*_spin_<10^-4^) and structural (*r* = -0.24, *P*_spin_<10^-4^; *r* = -0.23, *P*_spin_=10^-4^) epicenters, indicating that low connected regions in sensorimotor areas most closely resemble disease-related connectivity patterns. Similar correlation profiles were observed when assessing the same associations using MIND degree and gradients derived from SSD patients (see Supplementary Fig. 3).

### Baseline MIND alterations in SSD are co-localized with cortical hierarchy and SCZ epicenters

Significant differences in MIND degree and G1 were found between individuals with SSD and HC at baseline (Fig. 2A). A significant reduction in degree was observed in SSD in the left superior temporal and nucleus accumbens regions, along with the bilateral pallidum, whereas the lingual area showed significantly higher degree. Additionally, G1 was significantly increased in the bilateral lingual region and in the right cuneus, while a reduction was observed in the middle temporal area. However, no significant differences in G2 were observed at baseline. The effects of the remaining model covariates on MIND are shown in Supplementary Fig. 4.

We additionally hypothesized that these structural alterations in SSD would be co-localized with MIND features themselves (degree, G1, and G2). Consistent with this, we found that highly connected regions in HC exhibited stronger degree reductions in SSD (Fig. 2B; degree, *r* = -0.31, *P*_spin_<10^-4^). Furthermore, cortical gradient alterations were stronger and oppositely directed in regions located at the extremes of the gradients, indicating a compression along the gradient (G1, *r* = -0.27; G2, *r* = -0.55; *P*_spin_<10^-4^). For G2, this compression reflected a lower differentiation—or dedifferentiation—between sensorimotor and association regions. However, the ranges of G1 and G2 were not significantly associated with SSD diagnosis (see Supplementary Fig. 5). Collectively, these findings revealed that regions with higher MIND feature values showed stronger reductions of that feature in SSD. Consequently, degree, G1, and G2 of HC were accounted for using partial correlation when assessing the spatial co-localization of structural differences with the spatial brain maps. These analyses revealed that evolutionary expansion was negatively correlated with baseline disease-related alterations in degree, G1, and G2 in SSD (Fig. 2C; *r*_partial_=-0.34, *r*_partial_=-0.39, and *r*_partial_=-0.29, respectively; *P*_spin_<10^-4^), indicating that baseline reductions in similarity and gradients in SSD mainly occur in regions exhibiting greater evolutionary expansion. The functional gradient and the S-A ranking were also negatively correlated with degree (*r*_partial_=-0.16, *P*_spin_=0.006, and *r*_partial_=-0.32, *P*_spin_<10^-4^, respectively) and G1 (*r*_partial_=-0.18, *P*_spin_=0.002; *r*_partial_=-0.26, *P*_spin_<10^-4^), revealing that these reductions are more specifically located in higher-order regions. On the contrary, a positive association was found for both functional (*r*_partial_=0.31; *r*_partial_=0.34, *P*_spin_<10^-4^) and structural (*r*_partial_=0.15, *P*_spin_=0.006; *r*_partial_=0.22, *P*_spin_<10^-4^) epicenters in schizophrenia, suggesting that decreases in degree and G1 most closely resemble disease-related morphological alteration patterns.

### MIND degree in SSD is associated with treatment duration and medication, and co-localized with hierarchy and SCZ epicenters

Although no significant associations were found in the longitudinal analyses between the main effect of diagnosis and degree, both treatment duration and medication were associated with MIND degree (Fig. 3A; see *Source Data* for *t*-values). The main effect of *time* was negatively associated with degree in the frontal, parietal, and temporal regions, along with the nucleus accumbens and the pallidum, indicating lower connectivity over time in these regions. Conversely, the occipital area, the thalamus, and the putamen exhibited a positive association, indicating an increase of degree over time. Interestingly, this effect turned out to be largely the opposite for individuals with SSD, who showed a positive *diagnosis:time* interaction, suggesting that the effect of time on MIND degree was attenuated in SSD. Consistent with this finding, the effect of *medication* also showed a positive association, indicating that the use of antipsychotics may contribute to a higher MIND degree. The longitudinal effects of the remaining model covariates on MIND degree are shown in Supplementary Fig. 6.

**Fig. 3.**
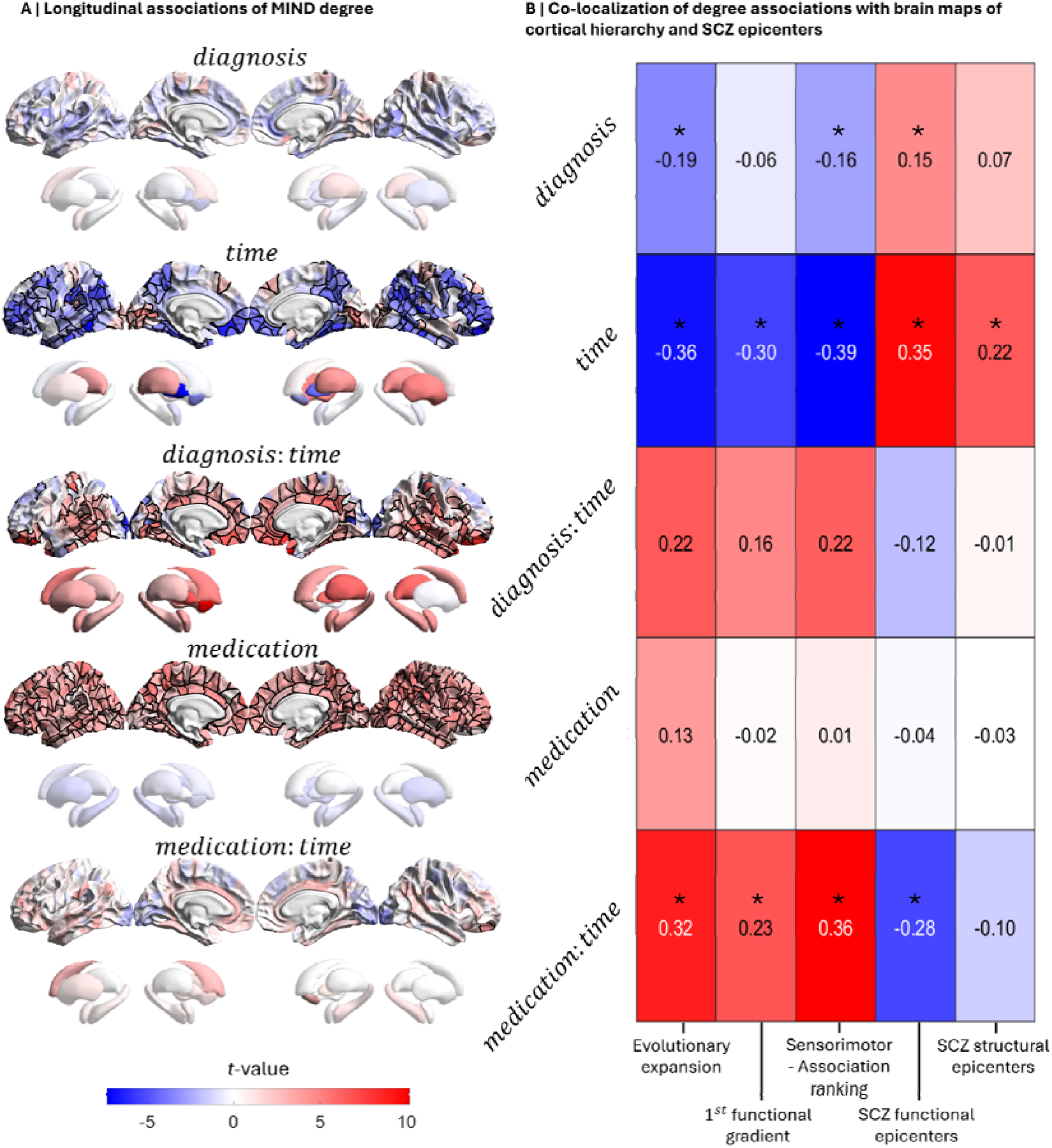
Longitudinal effects of diagnosis, time, and medication on MIND degree in SSD associated with hierarchical organization and SCZ epicenters. (**A**) Regional *t*-values representing the contribution of each variable to the LMM assessing the longitudinal relationship between MIND degree and covariates in SSD (FDR-corrected, *P*□<□0.05). (**B**) Regional co-localization (two-sided test) of the longitudinal *t*-values with brain maps of cortical hierarchy and SCZ epicenters, while accounting for regional brain connectivity. Asterisks (*) indicate significant correlations (FDR-corrected across features; *P*_spin_□<□0.05). MIND, morphometric inverse divergence; SCZ, schizophrenia.

Evolutionary expansion and the S-A axis were negatively co-localized with the effect of *diagnosis* (Fig. 3B; *r*_partial_=-0.19, *P*_spin_=0.008, and *r*_partial_=-0.16, *P*_spin_=0.038, respectively), indicating that cortical regions with reduced degree in SSD are higher-order association areas that present a larger surface area expansion in humans compared with macaques. In addition to these features, the functional gradient was also negatively associated with *time* (*r*_partial_=-0.36; *r*_partial_=-0.30; *r*_partial_=-0.39; *P*_spin_<10^-4^), revealing that regions with the strongest degree reductions over time are higher-order association transmodal (DMN) areas with greater evolutionary expansion. On the other hand, the functional epicenters in SCZ were positively associated with *diagnosis* (*r*_partial_=0.15, *P*_spin_=0.038) and *time* (*r*_partial_=0.35, *P*_spin_<10^-4^), while structural epicenters were only associated with *time* (*r*_partial_=0.22; *P*_spin_=0.001), indicating that regions whose connectivity most closely resembled disease-related connectivity alteration patterns exhibited higher connectivity in SSD and strongest degree increase over time. The model did not reveal a significant effect of the *medication:time* interaction; however the resulting *t*-value map was significantly co-localized with hierarchy and the functional SCZ epicenters.

### MIND gradients are associated with treatment duration, and co-localized with evolutionary expansion and SCZ epicenters

Although structural gradients were not significantly associated in the longitudinal analyses with the main effect of diagnosis, both treatment duration and its interaction with medication were associated with MIND gradients (Fig. 4A; see *Source Data* for *t*-values). The main effect of *time* on G1 showed widespread positive and negative associations, whereas its effect on G2 was more localized. Specifically, G2 decreased over time in frontal, cingulate, and temporal regions, while increasing in parietal and occipital regions, collectively reflecting a compression of the S-A ranking—that is, a reduction in hierarchical differences between somatosensory and association regions (see Supplementary Fig. 7 for *r*-values representing gradient compression). Accordingly, the ranges of G1 and G2 were negatively associated with the main effect of *time* (G1 range, *t* = -4.45, *P*<10^-5^; G2 range *t* = -6.28, *P*<10^-9^), and further compressed in SSD (*diagnosis:time;* G1 range, *t* = -2.47, *P* = 0.01; G2 range *t* = - 2.98, *P* = 0.003). Regarding the effect of time within SSD (*diagnosis:time*) on gradients, frontal and parietal regions showed negative associations in G2, whereas the cingulate cortex showed positive associations in both G1 and G2, suggesting region-specific reorganization and progression within the cortical hierarchy over the course of antipsychotic treatment. Unlike MIND degree, the effect of *medication* on connectivity gradients barely showed any significant association; instead, a *medication:time* interaction emerged, indicating that the effect of time in highly medicated patients partially reversed the main effect of *time*. The longitudinal effects of the remaining model covariates on MIND gradients are shown in Supplementary Fig. 8.

**Fig. 4.**
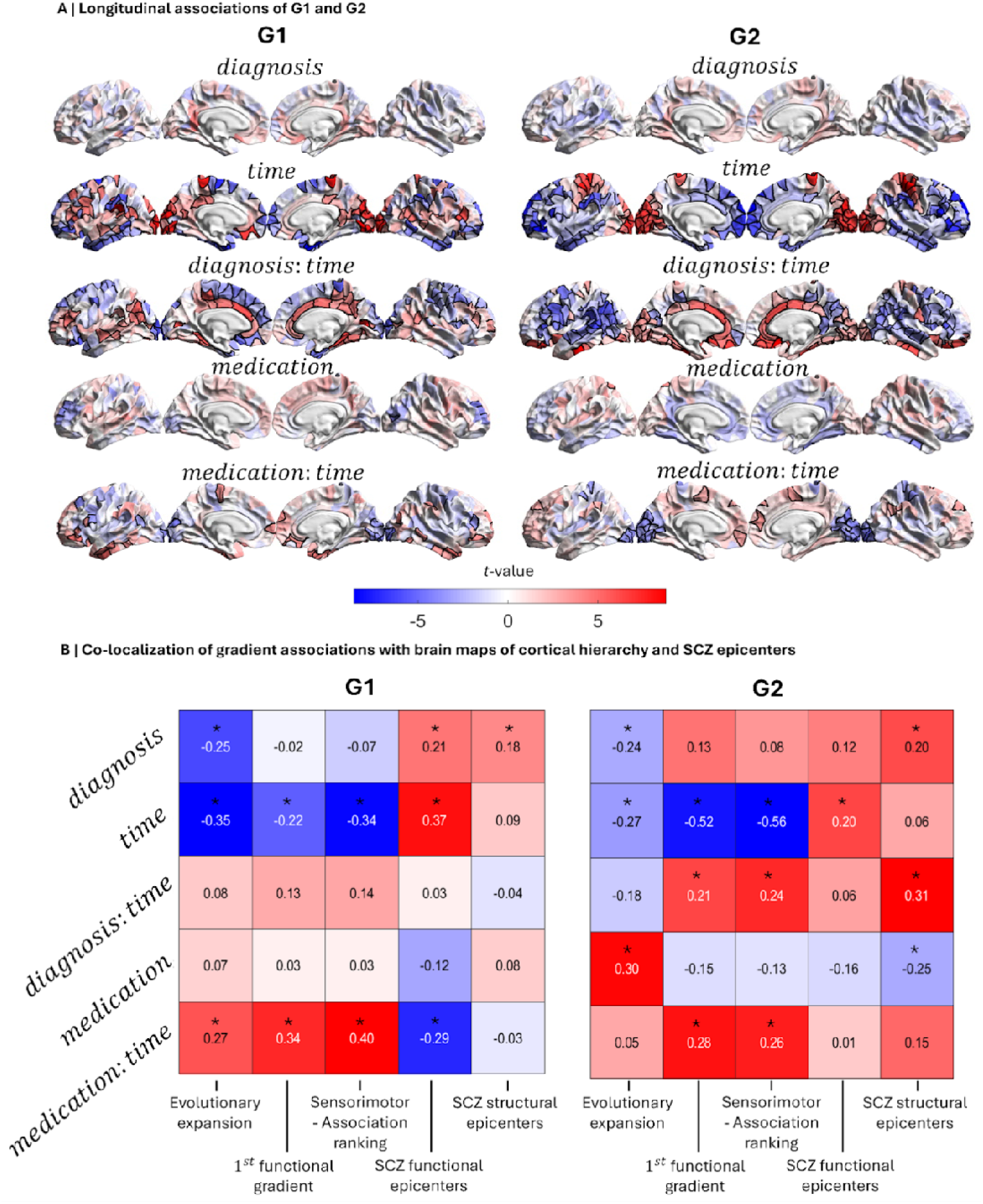
Longitudinal effects of diagnosis, time, and medication on MIND gradients in SSD associated with hierarchical organization and SCZ epicenters. (**A**) Regional *t*-values representing the contribution of each variable to the LMM assessing the longitudinal relationship between MIND gradients and covariates in SSD (FDR-corrected, *P* <□0.05). (**B**) Regional co-localization (two-sided test) of the longitudinal *t*-values with brain maps of cortical hierarchy and SCZ epicenters, while accounting for regional gradients. Asterisks (*) indicate significant correlations (FDR-corrected across features; *P*_spin_□<□0.05). G1, first gradient; G2, second gradient; SCZ, schizophrenia.

Subsequently, we found that the effect of *diagnosis* on G1 and G2 was negatively co-localized with evolutionary expansion (Fig. 4B; *r*_partial_=-0.25, *P*_spin_<10^-4^, and *r*_partial_=-0.24, *P*_spin_=0.005, respectively), indicating that regions with reduced gradients in SSD are primary regions with greater cortical expansion. Moreover, evolutionary expansion, the functional gradient, and the S-A axis were negatively co-localized with *time* (G1: *r*_partial_=-0.35, *P*_spin_<10^-4^; *r*_partial_=-0.22, *P*_spin_=0.03; and *r*_partial_=-0.34, *P*_spin_<10^-4^, respectively; G2: *r*_partial_=-0.27, *P*_spin_=2·10^-4^; *r*_partial_=-0.52, *P*_spin_<10^-4^; and *r*_partial_=-0.56, *P*_spin_<10^-4^, respectively), suggesting that the regions with the strongest gradient reductions over time are higher-order association DMN areas with greater evolutionary expansion in humans. Conversely, this effect was partially reversed in G2 of SSD individuals for the functional gradient and the S-A axis (*diagnosis:time; r*_partial_=0.21, *P*_spin_=0.019; and *r*_partial_=0.24, *P*_spin_=0.01). The effect of *medication* on G2 preferentially involved regions with greater evolutionary expansion (*r*_partial_=0.30, *P*_spin_=0.002). In addition, consistent with the opposite effect observed between the main effect of *time* and the *medication:time* interaction, all associations between *medication:time* and evolutionary expansion (G1: *r*_partial_=0.27, *P*_spin_=0.002), the functional gradient (G1: *r*_partial_=0.34, *P*_spin_<10^-4^; G2: *r*_partial_=0.28, *P*_spin_=0.007), and the S-A axis (G1: *r*_partial_=0.40, *P*_spin_<10^-4^; G2: *r*_partial_=0.26, *P*_spin_=0.02) were reversed and become positive.

Additionally, the functional epicenters in SCZ showed significant associations with *diagnosis* (G1: *r*_partial_=0.21, *P*_spin_=0.001), *time* (G1: *r*_partial_=0.37, *P*_spin_<10^-4^; G2: *r*_partial_=0.20, *P*_spin_=0.005), and *medication:time* (G1: *r*_partial_=-0.29, *P*_spin_<10^-4^), indicating that functional epicenters exhibited higher gradients in SSD and strongest gradient increase over time, except in patients prescribed with higher doses. Structural epicenters also showed associations with *diagnosis* (G1: *r*_partial_=0.18, *P*_spin_=0.01; G2: *r*_partial_=0.20, *P*_spin_=0.03) and *diagnosis:time* (G2: *r*_partial_=0.31, *P*_spin_=0.008), indicating that structural epicenters exhibited higher gradients and strongest gradient increase over time in SSD.

### MIND is associated with the baseline and long-term course of psychiatric symptoms

The impact of baseline MIND degree and gradients on psychiatric symptomatology revealed that MIND degree was negatively associated with symptoms in the frontal, temporal, and rostral anterior cingulate regions, indicating that reduced connectivity relates to more severe outcomes (Fig. 5A; *degree*; see *Source Data* for *t*-values). However, the degree of subcortical structures, including the left amygdala and nucleus accumbens, showed a positive association. Similarly, gradients exhibited both positive and negative associations with BPRS that were broadly distributed across the cortex (*G1* and *G2*).

**Fig. 5.**
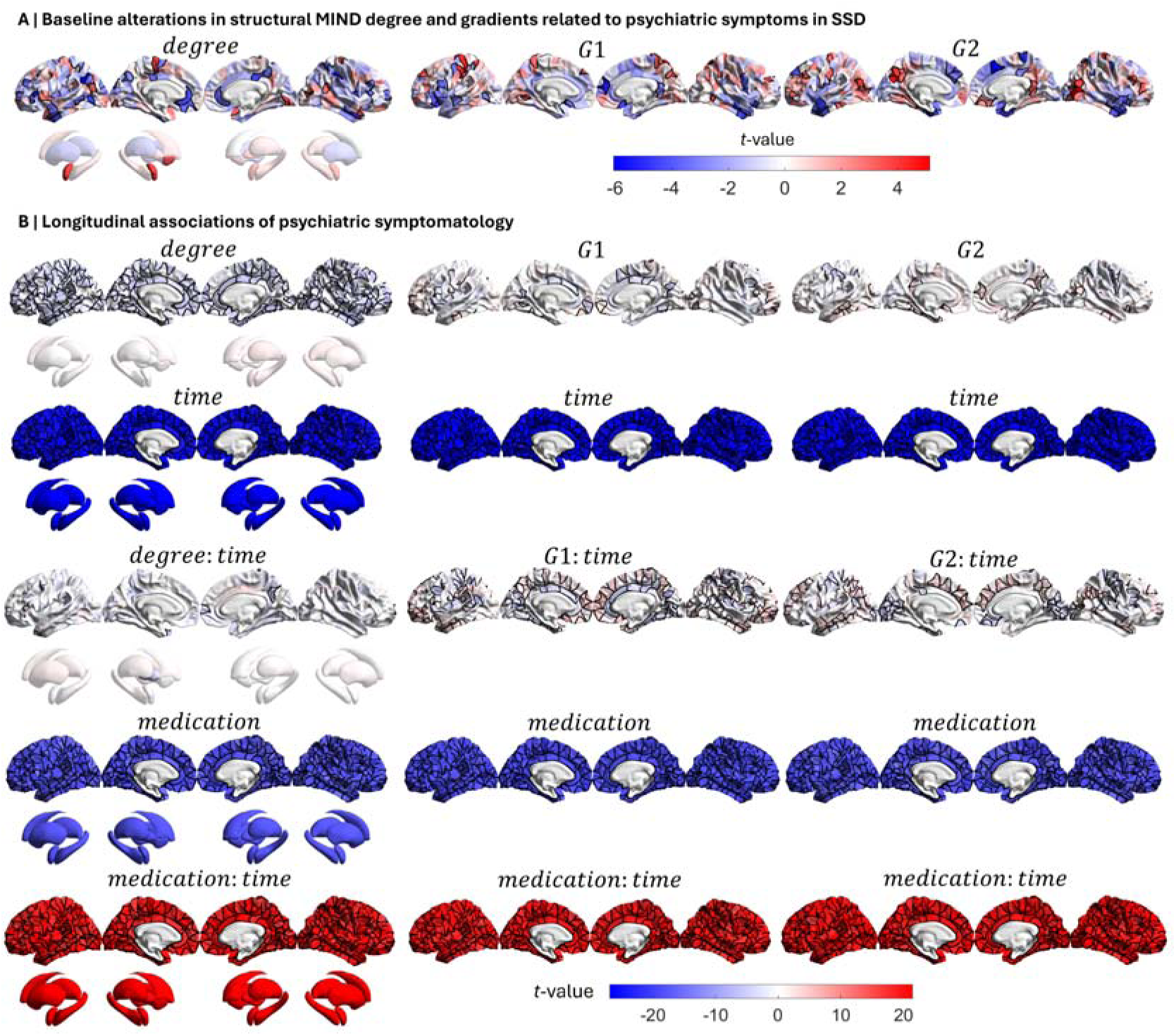
Baseline and longitudinal associations between psychiatric symptoms and brain similarity in SSD. (**A**) Regional *t*-values representing the contribution of each variable to the GLM assessing the baseline relationship between psychiatric symptoms—assessed by the Brief Psychiatric Rating Scale—and MIND (degree or gradients) in SSD, adjusting for covariates (from left to right: *degree, G1,* and *G2*; FDR-corrected, *P*□<□0.05). (**B**) Regional *t*-values representing the contribution of each variable to the GLMM assessing the longitudinal relationship between psychiatric symptoms and MIND in SSD, adjusting for covariates (FDR-corrected, *P*□<□0.05). G1, first gradient; G2, second gradient; MIND, morphometric inverse divergence; SSD, schizophrenia spectrum disorders.

The longitudinal associations of MIND degree and gradients with psychiatric symptomatology revealed that cortical degree was widely negatively associated with BPRS (Fig. 5B; *degree*). Gradients, however, exhibited both positive and negative associations broadly distributed across the cortex (*G1* and *G2*). As expected, the effect of *time* was negatively associated with BPRS scores in all models, indicating a reduction in overall psychiatric symptom severity over the course of treatment. Although the *degree:time* interaction barely showed any significant association, the *gradient:time* effect captured several positive and negative associations. According to treatment duration, the effect of *medication* also exhibited a negative association, indicating that symptoms decreased with antipsychotic dose. However, this effect reversed for high doses maintained over time (*medication:time*). The baseline and longitudinal effects of the remaining model covariates on psychiatric symptoms are shown in Supplementary Figs. 9 and 10, respectively.

## Discussion

In the present work, we investigated brain structural similarity by estimating cortical and subcortical Morphometric INverse Divergence (MIND) networks in healthy controls (HC) and individuals with schizophrenia spectrum disorders (SSD). We first examined the role of MIND-derived features (degree and gradients) in relation to SSD diagnosis, revealing that MIND was longitudinally associated with treatment duration and medication in SSD. Subsequently, we explored the co-localization of these MIND associations with spatial maps of cortical hierarchy and schizophrenia (SCZ) epicenters, identifying region-specific vulnerability to disease. Finally, we assessed the impact of baseline and longitudinal MIND on psychiatric symptoms, revealing that symptom improvement was associated with higher structural similarity and gradient changes over time.

Recent morphometric studies in schizophrenia have investigated individual patterns assessing structural similarity across regions, revealing network-level dysconnectivity and disruptions among anatomically connected brain regions [31, 42, 43]. Connectome-derived structural gradients are also being increasingly employed to characterize hierarchical brain organization and its role in neuropsychiatric disorders [27, 28]. In this study, we explored MIND connectivity by deriving the first (G1) and second (G2) structural gradients, with G2 capturing the sensorimotor-to-association (S-A) axis. This hierarchical representation of G2 aligned with the first connectome-derived functional gradient in both humans and the macaque monkey, which differentiates regions involved in primary sensorimotor functions from transmodal regions corresponding to the default mode network (DMN) [24]. Here, we reported significant differences in degree and gradients between SSD and HC individuals. In particular, a significant reduction in MIND degree was found at baseline in the temporal lobe, as well as in the nucleus accumbens and pallidum regions. This finding aligns with the dysconnectivity hypothesis of schizophrenia, which suggests a descending modulation of synaptic efficacy in hierarchically subordinate structures [44]. These reductions in MIND degree were observed in highly connected regions, consistent with previous studies showing that hub regions are particularly vulnerable to disease-related reductions in both functional and structural brain networks [33, 45]. Furthermore, we observed an increase in G1 in the occipital lobe, consistent with previous findings reporting alterations in the principal gradient in individuals with bipolar disorder using the same approach [27]. Additionally, alterations in G1 and G2 were stronger and oppositely directed in regions located at the extremes of the gradients, indicating a gradient compression along the S-A axis [46–48]. This dedifferentiation, i.e. a reduction in regional differentiation, has been hypothesized to be linked to genes involved in synaptic signaling and neuronal development [47]. Together, these findings on MIND structure suggest that these affected regions are more structurally differentiated from each other and less interconnected in SSD compared to HC [42]. Nevertheless, the longitudinal analyses revealed that the effects of SSD diagnosis on MIND were related to treatment duration and medication. In particular, the temporal progression indicated a generalized reduction in cortical similarity and gradient ranges over time [26]. In SSD, the time-related decreases in both cortical and subcortical similarity were partially reversed, while the cortical gradient ranges exhibited further compression. This may be attributed to the relevant role of antipsychotics and the course of disease progression in shaping brain structure, as evidenced by previous longitudinal studies in schizophrenia [4, 9]. Consistently, medication was also associated with MIND degree, revealing an increase in inter-regional similarity with antipsychotic use, which aligns with reported improvements in white matter microstructural integrity and corticostriatal functional connectivity associated with antipsychotic exposure in first-episode psychosis and schizophrenia [49–51].

Structural alterations may initially emerge as epicenters in specific brain regions that are particularly vulnerable to disease, subsequently spreading across the rest of the brain [33], potentially disrupting connectivity and hierarchical organization. In our study, we found that MIND associations were co-localized with cortical hierarchy and SCZ epicenters both at baseline and longitudinally. Specifically, evolutionary expansion was associated with baseline and longitudinal changes in degree and gradient organization in SSD. The S-A ranking was also co-localized with baseline alterations in MIND degree and G1, aligning with the hierarchy-dependent functional dysconnectivity in schizophrenia [52]. Moreover, both the S-A ranking and the functional gradient were co-localized with longitudinal changes in G2 in individuals with SSD. These findings are consistent with previous studies in schizophrenia demonstrating reduced hierarchy [53] and preferential network-level disruptions in association regions [54], along with the rapid evolutionary cortical expansion of the DMN associated with high expression of genes involved in schizophrenia [55]. Furthermore, SCZ epicenters were co-localized with MIND degree and G1 at baseline in SSD, whereas the co-localization of G2 with structural epicenters was time-dependent in SSD, supporting the hypothesis that the underlying connectivity patterns facilitate the progression of structural alterations [56]. This finding supports emerging evidence exploring the role of excitatory-inhibitory neuronal imbalance in the preferential vulnerability of higher-order association networks [54, 57]. Altogether, the spatial co-localization of MIND associations with cortical hierarchy and SCZ epicenters reflects a region-specific pattern of vulnerability during SSD onset and progression.

Previous studies have demonstrated that structural connectivity alterations are associated with clinical symptomatology [58, 59], including studies using structural connectivity to predict psychopathological deficits [19]. Notably, functional connectome biomarkers have been shown to successfully predict positive symptoms in adolescent-onset schizophrenia [60]. Accordingly, we found that baseline MIND-symptom associations progressively changed during treatment duration. This finding supports the relevant role of disrupted connectivity and hierarchy to clinical symptomatology in SSD [43, 61–64]. Moreover, the effect of medication indicated that psychiatric symptoms decreased with increasing antipsychotic dosage, consistent with findings from a previous triple-blind, placebo-controlled study supporting the beneficial effects of antipsychotics in attenuating psychiatric symptomatology in first-episode psychosis [9]. However, in line with previous studies suggesting that high doses of antipsychotics are less effective for treating negative symptoms [65], we found that long-term exposure to high doses was related to more severe symptomatology in SSD.

The present findings must be interpreted with several considerations. First, this is a long-running study that inevitably experienced high drop-out rates, further increasing the risk of attrition bias. Second, since all patients were medicated and no untreated comparison group was included due to ethical considerations, we cannot disentangle the effects of medication from the natural course of the disorder. Third, higher medication doses were typically prescribed to individuals with more severe psychiatric symptoms; however, prescribing practices were also influenced by factors such as age at onset and treatment duration, complicating the disentanglement of their independent effects. Fourth, evolutionary expansion represented the surface area ratio between humans and macaques [66]; however, bridging the gap between humans and extant mammals requires reconstructing the cortical shape of their common ancestor [67].

In summary, we identified that cortical and subcortical MIND were longitudinally associated with treatment duration and medication in SSD. Additionally, these associations were co-localized with brain maps of cortical hierarchy and epicenters in schizophrenia, highlighting the relevant role of regional vulnerability in disease onset and progression. Moreover, psychiatric symptoms decreased as structural similarity increased, with gradients being related to treatment duration. Collectively, these findings contribute to advancing the understanding of how hierarchical and evolutionary processes of brain organization, together with treatment duration and medication, shape structural similarity and clinical symptoms throughout the course of SSD.

## Data availability

The brain maps of cortical hierarchy were obtained using *neuromaps* toolbox [40], available at https://github.com/netneurolab/neuromaps. The brain maps of functional and structural epicenters in SCZ, available at https://enigma-toolbox.readthedocs.io/en/latest/pages/07.epicenter/index.html, were obtained from an ENIGMA study [33].

## Code availability

All code and non-clinical data used to perform the analyses can be found at https://github.com/NeuroimagingBrainNetworks/HierarchyLongitudinalMINDGradientsPsychosis/ [68]. The ENIGMA-Shape Analysis pipeline employed to estimate surface meshes for subcortical structures can be downloaded at https://enigma.ini.usc.edu/ongoing/enigma-shape-analysis/. The code used for computing subcortical MIND networks was derived from Cardoso Saraiva *et al.* [37], and can be found at https://github.com/NeuroimagingBrainNetworks/HierarchyLongitudinalMINDGradientsPsychosis/ [68]. The code used for computing cortical MIND networks can be found at https://github.com/isebenius/MIND.

## Data Availability

The brain maps of cortical hierarchy were obtained using neuromaps toolbox, available at https://github.com/netneurolab/neuromaps. The brain maps of functional and structural epicenters in SCZ, available at https://enigma-toolbox.readthedocs.io/en/latest/pages/07.epicenter/index.html, were obtained from an ENIGMA study.

https://github.com/NeuroimagingBrainNetworks/HierarchyLongitudinalMINDGradientsPsychosis

## Acknowledgments

RRG is funded by the *Plan de Generación de Conocimiento* from the Spanish Ministry of Science (PID2021-122853OA-I00), and ERANET Neuron JTC 2023 (ERP-2023-23684211). Both RRG and NGS are funded by the *Plan de Consolidación* (CNS2023-143647). JS is funded by the Psychosis Immune Mechanism Stratified Medicine Study (PIMS), UK Medical Research Council, MR/S037675/1. All research at the Department of Psychiatry at the University of Cambridge is supported by the NIHR Cambridge Biomedical Research Centre (NIHR203312) and the NIHR Applied Research Collaboration East of England. The views expressed are those of the author(s) and not necessarily those of the NIHR or the Department of Health and Social Care.

## Author Contributions

N.G.S. performed data curation, methodological design, data analysis, and drafted the manuscript; R.A.I.B., I.S., L.C.S., P.S., C.A.M., C.G., P.S.Q., A.P., G.S., M.R.V., R.A.A., J.V.B., B.M., C.C., J.S., B.C.F., and R.R.G contributed to data acquisition, provided advice on data analysis, and participated in writing and editing the manuscript. R.R.G. also contributed to conceptualization and supervision of the work. All authors approved the submitted version of the manuscript.

## Competing interests

The authors declare no competing interests.

## Notes

### Competing Interest Statement

The authors have declared no competing interest.

### Summary of Updates

This version of the manuscript has been revised to upload the supplemetary material and supplementary data

## References

1. Wen K, Zhao Y, Gong Q, Zhu Z, Li Q, Pan N, et al. Cortical thickness abnormalities in patients with first episode psychosis: a meta-analysis of psychoradiologic studies and replication in an independent sample. Psychoradiology. 2021;1:185–198.

2. Muñoz-Caracuel M, Alemán-Morillo C, García-San-Martín N, Garrido-Torres N, Alemany-Navarro M, Bethlehem RAI, et al. Predicting clinical and functional trajectories in individuals with first-episode psychosis by baseline deviations in grey matter volume. The British Journal of Psychiatry. 2025:1–9.

3. Kottaram A, Johnston LA, Tian Y, Ganella EP, Laskaris L, Cocchi L, et al. Predicting individual improvement in schizophrenia symptom severity at 1-year follow-up: Comparison of connectomic, structural, and clinical predictors. Human Brain Mapping. 2020;41:3342–3357.

4. Alemán-Morillo C, García-San-Martín N, Bethlehem RAI, Dorfschmidt L, Alemany-Navarro M, Segura P, et al. Medication and atypical brain maturation in psychosis associated with long-term cognitive decline and symptom progression. The British Journal of Psychiatry. 2025:1–11.

5. Zhao Y, Zhang Q, Shah C, Li Q, Sweeney JA, Li F, et al. Cortical Thickness Abnormalities at Different Stages of the Illness Course in Schizophrenia: A Systematic Review and Meta-analysis. JAMA Psychiatry. 2022;79:560–570.

6. Andreasen NC. The lifetime trajectory of schizophrenia and the concept of neurodevelopment. Dialogues in Clinical Neuroscience. 2010;12:409–415.

7. Dietsche B, Kircher T, Falkenberg I. Structural brain changes in schizophrenia at different stages of the illness: A selective review of longitudinal magnetic resonance imaging studies. Aust N Z J Psychiatry. 2017;51:500–508.

8. Pantelis C, Velakoulis D, McGorry PD, Wood SJ, Suckling J, Phillips LJ, et al. Neuroanatomical abnormalities before and after onset of psychosis: a cross-sectional and longitudinal MRI comparison. The Lancet. 2003;361:281–288.

9. Chopra S, Fornito A, Francey SM, O’Donoghue B, Cropley V, Nelson B, et al. Differentiating the effect of antipsychotic medication and illness on brain volume reductions in first-episode psychosis: A Longitudinal, Randomised, Triple-blind, Placebo-controlled MRI Study. Neuropsychopharmacol. 2021;46:1494–1501.

10. Chopra S, Holmes A, Segal A, Zhang X-H, Francey SM, O’Donoghue B, et al. Differential effects of illness and antipsychotics on cortical thinning in first episode psychosis: A randomised placebo-controlled MRI study. 2025:2025.05.08.25327204.

11. de Castro-Manglano P, Mechelli A, Soutullo C, Gimenez-Amaya J, Ortuño F, McGuire P. Longitudinal changes in brain structure following the first episode of psychosis. Psychiatry Research: Neuroimaging. 2011;191:166–173.

12. Chopra S, Segal A, Oldham S, Holmes A, Sabaroedin K, Orchard ER, et al. Network-Based Spreading of Gray Matter Changes Across Different Stages of Psychosis. JAMA Psychiatry. 2023;80:1246–1257.

13. Shafiei G, Markello RD, Makowski C, Talpalaru A, Kirschner M, Devenyi GA, et al. Spatial Patterning of Tissue Volume Loss in Schizophrenia Reflects Brain Network Architecture. Biological Psychiatry. 2020;87:727–735.

14. Sebenius I, Seidlitz J, Warrier V, Bethlehem RAI, Alexander-Bloch A, Mallard TT, et al. Robust estimation of cortical similarity networks from brain MRI. Nat Neurosci. 2023;26:1461–1471.

15. Cardoso Saraiva L, Sato JR, Sebenius I, Dzinalija N, del Río-Torné C, Godinho F, et al. Regional, functional and transcriptomic decoding of multidimensional brain structure alterations in obsessive-compulsive disorder. Nat Commun. 2026. 24 June 2026. 10.1038/s41467-026-74153-2.

16. King DJ, Wood AG. Clinically feasible brain morphometric similarity network construction approaches with restricted magnetic resonance imaging acquisitions. Netw Neurosci. 2020;4:274–291.

17. Yao G, Luo J, Zou T, Li J, Hu S, Yang L, et al. Transcriptional patterns of the cortical Morphometric Inverse Divergence in first-episode, treatment-naïve early-onset schizophrenia. NeuroImage. 2024;285:120493.

18. Montagnese M, Ebneabbasi A, García-San-Martín N, Pecci-Terroba C, Romero-García R, Morgan SE, et al. Structural similarity networks reveal brain vulnerability in dementia. Alzheimer’s & Dementia. 2025;21:e70973.

19. Sun Y, Zhang Z, Kakkos I, Matsopoulos GK, Yuan J, Suckling J, et al. Inferring the Individual Psychopathologic Deficits With Structural Connectivity in a Longitudinal Cohort of Schizophrenia. IEEE Journal of Biomedical and Health Informatics. 2022;26:2536–2546.

20. Bullmore E, Sporns O. Complex brain networks: graph theoretical analysis of structural and functional systems. Nat Rev Neurosci. 2009;10:186–198.

21. Smith SM, Fox PT, Miller KL, Glahn DC, Fox PM, Mackay CE, et al. Correspondence of the brain’s functional architecture during activation and rest. Proc Natl Acad Sci U S A. 2009;106:13040–13045.

22. Cole MW, Yarkoni T, Repovš G, Anticevic A, Braver TS. Global Connectivity of Prefrontal Cortex Predicts Cognitive Control and Intelligence. J Neurosci. 2012;32:8988–8999.

23. Greicius MD, Krasnow B, Reiss AL, Menon V. Functional connectivity in the resting brain: A network analysis of the default mode hypothesis. Proceedings of the National Academy of Sciences. 2003;100:253–258.

24. Margulies DS, Ghosh SS, Goulas A, Falkiewicz M, Huntenburg JM, Langs G, et al. Situating the default-mode network along a principal gradient of macroscale cortical organization. Proceedings of the National Academy of Sciences. 2016;113:12574–12579.

25. Goodale MA, Milner AD. Separate visual pathways for perception and action. Trends in Neurosciences. 1992;15:20–25.

26. Taylor HP, Huynh KM, Thung K-H, Lin G, Lyu W, Lin W, et al. Functional hierarchy of the human neocortex across the lifespan. Nature. 2026:1–10.

27. Wang R, Xu J, Li F, Huang X, Xia C, Lui S, et al. Cortical morphometric gradients reveal molecular and cognitive underpinnings of bipolar disorder. Psychological Medicine. 2025;55:e383.

28. Han Y, Wang X, Cheng S, Yan P, Chen Y, Kang N, et al. Cortical morphometric similarity gradient in schizophrenia and its association with transcriptional profiles and clinical phenotype. Psychological Medicine. 2025;55:e97.

29. van den Heuvel MP, Scholtens LH, de Lange SC, Pijnenburg R, Cahn W, van Haren NEM, et al. Evolutionary modifications in human brain connectivity associated with schizophrenia. Brain. 2019;142:3991–4002.

30. Radua J, Borgwardt S, Crescini A, Mataix-Cols D, Meyer-Lindenberg A, McGuire PK, et al. Multimodal meta-analysis of structural and functional brain changes in first episode psychosis and the effects of antipsychotic medication. Neuroscience & Biobehavioral Reviews. 2012;36:2325–2333.

31. García-San-Martín N, Bethlehem RA, Segura P, Mihalik A, Seidlitz J, Sebenius I, et al. Reduced brain structural similarity is associated with maturation, neurobiological features, and clinical status in schizophrenia. Nat Commun. 2025;16:8745.

32. García-San-Martín N, Bethlehem RAI, Mihalik A, Seidlitz J, Sebenius I, Alemán-Morillo C, et al. Molecular and micro-architectural mapping of gray matter alterations in psychosis. Mol Psychiatry. 2025;30:1287–1296.

33. Georgiadis F, Larivière S, Glahn D, Hong LE, Kochunov P, Mowry B, et al. Connectome architecture shapes large-scale cortical alterations in schizophrenia: a worldwide ENIGMA study. Mol Psychiatry. 2024;29:1869–1881.

34. Crespo-Facorro B, Pérez-Iglesias R, Ramirez-Bonilla M, Martínez-García O, LLorca J, Vázquez-Barquero JL. A Practical Clinical Trial Comparing Haloperidol, Risperidone, and Olanzapine for the Acute Treatment of First-Episode Nonaffective Psychosis. J Clin Psychiatry. 2006;67:1511–1521.

35. Reuter M, Schmansky NJ, Rosas HD, Fischl B. Within-subject template estimation for unbiased longitudinal image analysis. Neuroimage. 2012;61:1402–1418.

36. Romero-Garcia R, Atienza M, Clemmensen LH, Cantero JL. Effects of network resolution on topological properties of human neocortex. NeuroImage. 2012;59:3522–3532.

37. Saraiva LC, Sato J, Sebenius I, Dzinalija N, Río-Torné C del, Godinho F, et al. Regional, functional and transcriptomic decoding of multidimensional brain structure alterations in obsessive-compulsive disorder. 2025.

38. Gardner M, Shinohara RT, Bethlehem RAI, Romero-Garcia R, Warrier V, Dorfschmidt L, et al. ComBatLS: A Location- and Scale-Preserving Method for Multi-Site Image Harmonization. Human Brain Mapping. 2025;46:e70197.

39. Vos de Wael R, Benkarim O, Paquola C, Lariviere S, Royer J, Tavakol S, et al. BrainSpace: a toolbox for the analysis of macroscale gradients in neuroimaging and connectomics datasets. Commun Biol. 2020;3:103.

40. Markello RD, Hansen JY, Liu Z-Q, Bazinet V, Shafiei G, Suárez LE, et al. neuromaps: structural and functional interpretation of brain maps. Nat Methods. 2022;19:1472–1479.

41. Ayesa-Arriola R, Ortíz-García De La Foz V, Martínez-García O, Setién-Suero E, Ramírez ML, Suárez-Pinilla P, et al. Dissecting the functional outcomes of first episode schizophrenia spectrum disorders: a 10-year follow-up study in the PAFIP cohort. Psychol Med. 2021;51:264–277.

42. Morgan SE, Seidlitz J, Whitaker KJ, Romero-Garcia R, Clifton NE, Scarpazza C, et al. Cortical patterning of abnormal morphometric similarity in psychosis is associated with brain expression of schizophrenia-related genes. Proc Natl Acad Sci U S A. 2019;116:9604–9609.

43. Li P, Jing R-X, Zhao R-J, Shi L, Sun H-Q, Ding Z, et al. Association between functional and structural connectivity of the corticostriatal network in people with schizophrenia and unaffected first-degree relatives. Journal of Psychiatry and Neuroscience. 2020;45:395–405.

44. Friston K, Brown HR, Siemerkus J, Stephan KE. The dysconnection hypothesis (2016). Schizophr Res. 2016;176:83–94.

45. Crossley NA, Mechelli A, Scott J, Carletti F, Fox PT, McGuire P, et al. The hubs of the human connectome are generally implicated in the anatomy of brain disorders. Brain. 2014;137:2382–2395.

46. Dong D, Yao D, Wang Y, Hong S-J, Genon S, Xin F, et al. Compressed sensorimotor-to-transmodal hierarchical organization in schizophrenia. Psychological Medicine. 2023;53:771–784.

47. Xiang J, Ma C, Chen X, Cheng C. Investigating Connectivity Gradients in Schizophrenia: Integrating Functional, Structural, and Genetic Perspectives. Brain Sci. 2025;15:179.

48. Hong S-J, Vos de Wael R, Bethlehem RAI, Lariviere S, Paquola C, Valk SL, et al. Atypical functional connectome hierarchy in autism. Nat Commun. 2019;10:1022.

49. Reis Marques T, Taylor H, Chaddock C, Dell’Acqua F, Handley R, Reinders AATS, et al. White matter integrity as a predictor of response to treatment in first episode psychosis. Brain. 2014;137:172–182.

50. Blazer A, Chengappa KNR, Foran W, Parr AC, Kahn CE, Luna B, et al. Changes in corticostriatal connectivity and striatal tissue iron associated with efficacy of clozapine for treatment□resistant schizophrenia. Psychopharmacology (Berl). 2022;239:2503–2514.

51. Sarpal DK, Robinson DG, Lencz T, Argyelan M, Ikuta T, Karlsgodt K, et al. Antipsychotic Treatment and Functional Connectivity of the Striatum: a Prospective Controlled Study in First-Episode Schizophrenia. JAMA Psychiatry. 2015;72:5–13.

52. David I, Sasai S, De Paiva FB, Boly M, Tononi G, Albantakis L. Distance- and hierarchy-dependent functional dysconnectivity in schizophrenia and its association with cortical microstructure. NeuroImage: Clinical. 2026;49:103958.

53. Bassett DS, Bullmore E, Verchinski BA, Mattay VS, Weinberger DR, Meyer-Lindenberg A. Hierarchical Organization of Human Cortical Networks in Health and Schizophrenia. J Neurosci. 2008;28:9239–9248.

54. Yang GJ, Murray JD, Wang X-J, Glahn DC, Pearlson GD, Repovs G, et al. Functional hierarchy underlies preferential connectivity disturbances in schizophrenia. Proceedings of the National Academy of Sciences. 2016;113:E219–E228.

55. Wei Y, de Lange SC, Scholtens LH, Watanabe K, Ardesch DJ, Jansen PR, et al. Genetic mapping and evolutionary analysis of human-expanded cognitive networks. Nat Commun. 2019;10:4839.

56. Jiang Y, Palaniyappan L, Luo C, Chang X, Zhang J, Tang Y, et al. Neuroimaging epicenters as potential sites of onset of the neuroanatomical pathology in schizophrenia. Science Advances. 2024;10:eadk6063.

57. Anticevic A, Lisman J. How Can Global Alteration of Excitation/Inhibition Balance Lead to the Local Dysfunctions That Underlie Schizophrenia? Biological Psychiatry. 2017;81:818–820.

58. Fornito A, Zalesky A, Pantelis C, Bullmore ET. Schizophrenia, neuroimaging and connectomics. NeuroImage. 2012;62:2296–2314.

59. van den Heuvel MP, Fornito A. Brain Networks in Schizophrenia. Neuropsychol Rev. 2014;24:32–48.

60. Fan Y-S, Li L, Peng Y, Li H, Guo J, Li M, et al. Individual-specific functional connectome biomarkers predict schizophrenia positive symptoms during adolescent brain maturation. Human Brain Mapping. 2021;42:1475–1484.

61. Holmes A, Levi PT, Chen Y-C, Chopra S, Aquino KM, Pang JC, et al. Disruptions of Hierarchical Cortical Organization in Early Psychosis and Schizophrenia. Biological Psychiatry: Cognitive Neuroscience and Neuroimaging. 2023;8:1240–1250.

62. Yang C, Zhang W, Liu J, Yao L, Bishop JR, Lencer R, et al. Disrupted subcortical functional connectome gradient in drug-naïve first-episode schizophrenia and the normalization effects after antipsychotic treatment. Neuropsychopharmacol. 2023;48:789–796.

63. Yao G, Luo J, Li J, Feng K, Liu P, Xu Y. Functional gradient dysfunction in drug-naïve first-episode schizophrenia and its correlation with specific transcriptional patterns and treatment predictions. Psychological Medicine. 2024;54:4106–4118.

64. Yang C, Zhang L, Liu J, Li K, Li S, Yang Z, et al. More Severe Brain Network Hierarchy Disorganization in Treatment-Naive Deficit Compared to Non-deficit Schizophrenia and Underlying Neurotransmitter Associations. Schizophr Bull. 2026;52:sbae231.

65. Oosthuizen P, Emsley R, Jadri Turner H, Keyter N. A randomized, controlled comparison of the efficacy and tolerability of low and high doses of haloperidol in the treatment of first-episode psychosis. Int J Neuropsychopharmacol. 2004;7:125–131.

66. Hill J, Inder T, Neil J, Dierker D, Harwell J, Van Essen D. Similar patterns of cortical expansion during human development and evolution. Proc Natl Acad Sci U S A. 2010;107:13135–13140.

67. Schwartz E, Nenning K-H, Heuer K, Jeffery N, Bertrand OC, Toro R, et al. Evolution of cortical geometry and its link to function, behaviour and ecology. Nat Commun. 2023;14:2252.

68. García-San-Martín N. NeuroimagingBrainNetworks/HierarchyLongitudinalMINDGradientsPsychosis: v1.0.0-alpha. 2026.

