## Supplemental Material for "Long-term morphometric similarity gradients relate to cortical hierarchy and psychiatric symptoms in schizophrenia"

**This PDF file includes:**

**Other Supplementary Information for this manuscript includes:**

Source Data (separate .xlsx file)

### Supplementary Subjects and Methods

#### **Dataset description**

Subjects from *Programa de Atención a las Fases Iniciales de Psicosis* (PAFIP) dataset were screened for the following criteria [1, 2]: (1) age 15 to 60 years; (2) DSM-IV criteria for a principal diagnosis of schizophreniform disorder, schizophrenia, schizoaffective disorder, brief reactive psychosis, schizotypal personality disorder, or psychosis not otherwise specified; (3) habitually living in the catchment area; (4) no prior treatment with antipsychotic medication or, if previously treated, a total lifetime of adequate antipsychotic treatment of less than 6 weeks. Patients were excluded for any of the following reasons: 1) met the DSM-IV criteria for drug dependence (except nicotine dependence), 2) met the DSM-IV criteria for mental retardation, or 3) had a history of neurological disease or head injury. There were no additional exclusion criteria for MRI except those specific to scanning logistics (e.g., claustrophobia, braces). The healthy volunteer group was recruited from the community through advertisements. They were required to have no current or previous psychiatric, mental retardation, neurological or general medical illnesses, including substance dependence and significant loss of consciousness, as determined by using an abbreviated version of the Comprehensive Assessment of Symptoms and History (CASH) [3]. They were selected to have a similar distribution in age, gender, and drug use history to the patient group. Clinical records and family interviews also confirmed the absence of psychosis in first-degree relatives. After a detailed description of the study, each subject gave written informed consent to participate in the study.

Participants who met criteria and provided written informed consent were enrolled in PAFIP. A total of 1293 images passed visual quality control (QC), corresponding to individuals who were scanned at baseline (*n*_SSD_ = 333; *n*_HC_ = 193) and underwent follow-up assessments at 1-year (*n*_SSD_ = 96; *n*_HC_ = 62), 3-years (*n*_SSD_ = 136; *n*_HC_ = 51), 5-years (*n*_SSD_ = 70; *n*_HC_ = 76), 10-years (*n*_SSD_ = 101; *n*_HC_ = 91), 15-years (*n*_SSD_ = 33; *n*_HC_ = 35), and 20-years (*n*_SSD_ = 8; *n*_HC_ = 8). However, after applying ENIGMA-Shape Analysis, subcortical structures underwent an additional QC, resulting in a total of *n*_SSD_ = 350 and *n*_HC_ = 193 participants with baseline data (*n*_SSD_ = 326; *n*_HC_ = 189) and subsequent follow-ups at 1-year (*n*_SSD_ = 95; *n*_HC_ = 62), 3-years (*n*_SSD_ = 134; *n*_HC_ = 51), 5-years (*n*_SSD_ = 69; *n*_HC_ = 74), 10-years (*n*_SSD_ = 98; *n*_HC_ = 91), 15-years (*n*_SSD_ = 31; *n*_HC_ = 35), and 20-years (*n*_SSD_ = 8; *n*_HC_ = 8).

#### **MRI acquisition**

MRI scans were obtained using a 1.5 T General Electric SIGNA System (GE Medical Systems, Milwaukee, WI) and a 3 T Philips Medical Systems MRI scanner (Achieva, Best, The Netherlands) at the Hospital Universitario Marqués de Valdecilla (HUMV). The parameters for 1.5 T were: TE = 5 ms, TR = 24 ms, NEX = 2, flip angle = 45° (for 32 follow-up images, the flip angle was set to 12° due to an unintended scanner protocol selection), FOV = 26×19.5 cm, slice thickness = 1.5 mm, and a matrix of 256×192, whereas for 3 T were: TE = 3.7 ms, TR = 8.2 ms, flip angle = 8°, acquisition matrix = 256×256, voxel size = 0.94x0.94x1 mm and 160 contiguous slices.

#### **Cortical and subcortical MIND estimation**

For each subject, a MIND network was constructed by computing between pairs of regions the Kullback-Leibler divergence of multiple MRI features. For the cortex these features included cortical thickness, mean curvature, sulcal depth, surface area, and gray matter volume. For the subcortex they included surface area and radial distance. Both Jacobian determinant—a proxy for surface area, representing the surface dilation ratio between the subcortical structure and a template at a corresponding vertex, that is, the surface deformation relative to a template—and radial distance—the distance between a surface vertex of the subcortical structure and its medial curve, defined as the curve that passes through the center of a three-dimensional structure—were derived and quality-controlled using ENIGMA-Shape Analysis (<https://enigma.ini.usc.edu/ongoing/enigma-shape-analysis/>).

#### **Cortical MIND gradients**

The first two structural connectivity gradients G1 and G2 were derived from cortical MIND networks (318x318) of each individual using gradient decomposition [4] with the following code parameters: GradientMaps (kernel=‘normalized angle’, approach=‘diffusion embedding’). This technique estimates a low-dimensional set of eigenvectors, or gradients, in which each gradient represents a different dimension of covariance in MIND, with a small number of gradients capturing most of the inter-regional similarity structure. In this reduced space, each gradient is anchored by regions with the strongest gradient values, indicating that the corresponding dimension effectively captures their hierarchical profiles [5]. In contrast, regions located near the origin (i.e., with low absolute gradient values) show minimal similarity to these anchor regions, suggesting that the gradient does not strongly capture their hierarchical profiles. Therefore, greater separation between the extremes of a gradient along its axis indicates stronger capture of hierarchical differentiation. To ensure comparability of gradients across individuals and eliminate directional randomness, Procrustes rotation [6] was subsequently applied to align each individual gradient map with a group-level gradient template derived from an average connectivity matrix encompassing both SSD and HC participants [7, 8].

#### **Baseline and longitudinal MIND associations in SSD**

To evaluate the impact of SSD diagnosis on brain similarity (degree and gradients) at baseline, multiple regression analyses were performed for each region, as shown in Eq. (1).

| Regional {degree, G1, or G2} ~ 1 + diagnosis + age at inclusion + sex + eTIV + Euler number | (1) |
| --- | --- |

where ‘*Regional{degree, G1, or G2}*’ denote the regional MIND degree or gradient, which were tested separately; ‘*diagnosis*’ is HC or SSD; ‘*age at inclusion*’ corresponds to the age at study enrolment; ‘*sex*’ is male or female; ‘*eTIV*’ is the estimated total intracranial volume for each individual; and ‘*Euler number*’ is the FreeSurfer’s Euler index, a proxy of motion-induced image artefacts.

The longitudinal effects of diagnosis, time, and medication on brain similarity were assessed using linear mixed modeling (LMM) as shown in Eq. (2).

| Regional {degree, G1, or G2} ~ 1 + diagnosis + age at inclusion + sex + eTIV + Euler number + time + diagnosis:time + medication + medication:time + protocol + (1\|subject) | (2) |
| --- | --- |

where, in addition to the aforementioned variables, ‘*time*’ represents the period since the enrolment for HC, or since treatment initiation for SSD; ‘*diagnosis:time*’ represents the interaction between diagnosis and time, i.e., the effect of treatment duration for SSD individuals; ‘*medication*’ corresponds to antipsychotic dose expressed in chlorpromazine equivalents, and was calculated according to the criterion described in [9, 10]; *‘medication:time*’ represents the interaction between medication and time, i.e., the effect of medication over time; ‘*protocol*’ is a binary variable that represents a change in the flip angle during MRI acquisition (see *MRI acquisition, parcellation, and volume extraction* section for details); and “*(1|subject)*” specifies a random intercept to account for subject-specific variability in overall offsets. For HC, ‘*medication*’ and its interaction term ‘*medication:time*’ were set to zero.

#### **Brain maps of cortical hierarchy and schizophrenia (SCZ) epicenters**

The map of evolutionary expansion represents the surface area ratio between humans and macaques [11]. The functional gradient corresponds to the first gradient of functional connectivity derived from diffusion map embedding [12]. The sensorimotor-association (S-A) regional ranking ranges from lower-order sensorimotor to higher-order associative functions, and was conformed by cortical features derived from remarkably varied data types including neuroimaging, histology, transcriptomics, receptor autoradiography, and electrophysiology [13]. The functional and structural epicenters in SCZ represent regions whose connectivity most closely resemble disease-related alteration patterns, i.e., regions that are connected to the strongest disease-related regional alterations [14].

#### **Spin test**

The spin test projects brain regions to a sphere using spherical coordinates generated during cortical-surface extraction. These spherical projections of brain annotation maps are rotated to randomize the relationship between cortical attributes and annotations [15, 16]. Each coordinate is assigned to its nearest rotated counterpart, resulting in a map where spatial autocorrelation is preserved, while the correspondence between parcels and annotations is randomized. Parcels that were closest to the medial wall were assigned the value of the nearest neighboring parcel instead. This procedure was performed at parcel resolution, rather than the vertex resolution, to prevent data upsampling, and it was repeated 10,000 times to generate a parcellation-specific rotation matrix (available at <https://github.com/frantisekvasa/rotate_parcellation>).

### Supplemental Figures

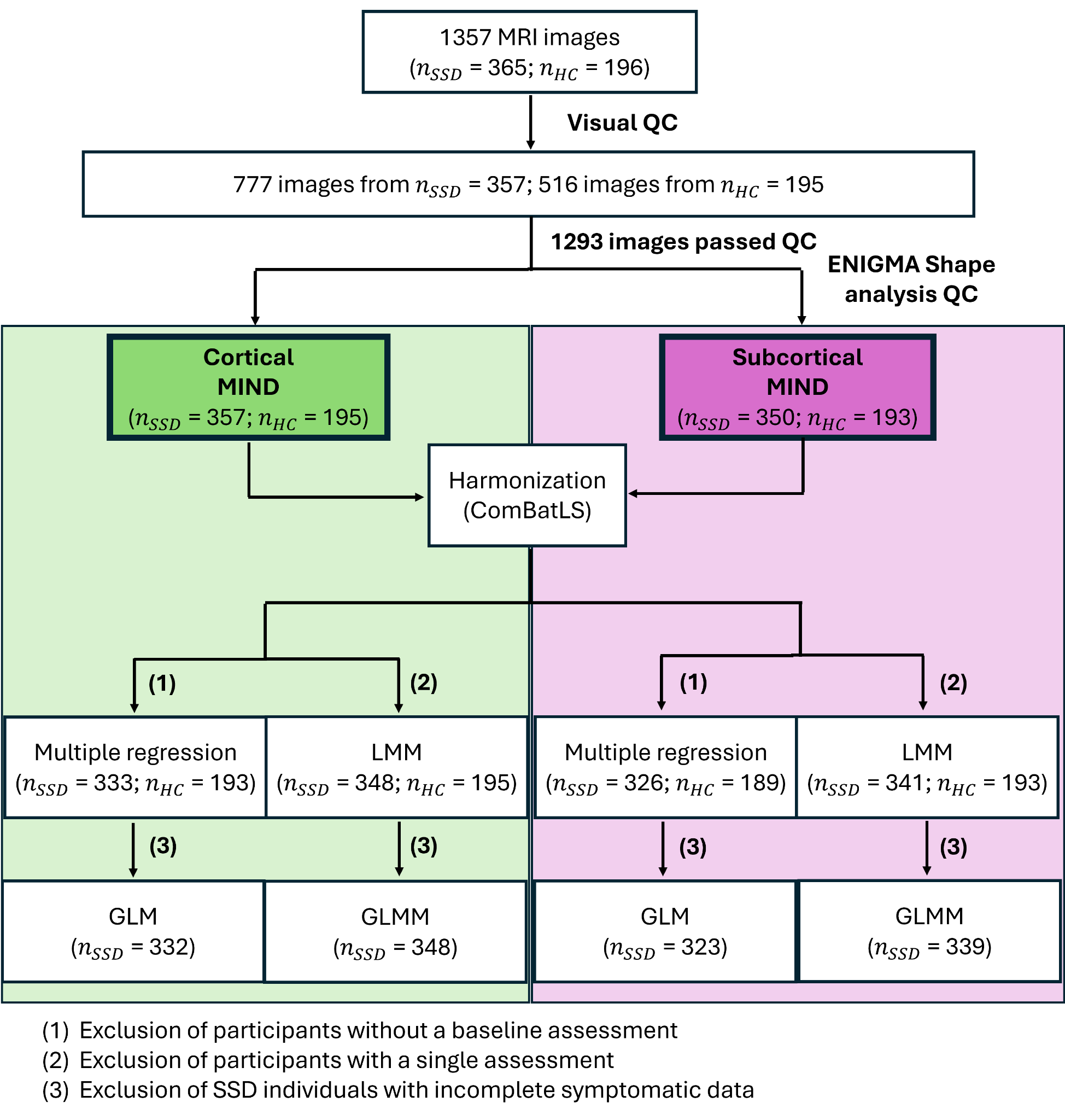

**Supplementary Fig. 1 Flow diagram of study participants.**

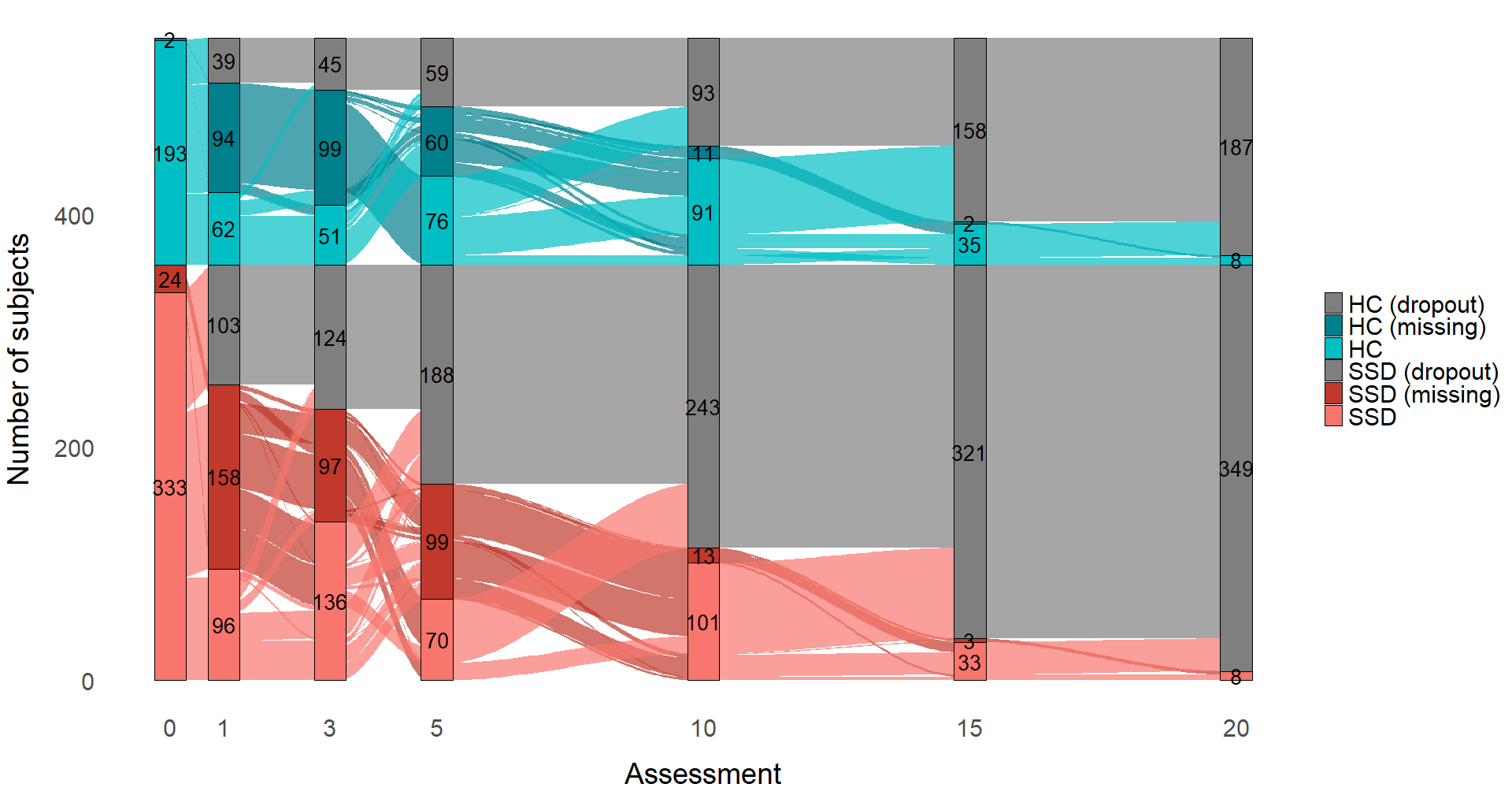

**Supplementary Fig. 2 Alluvial diagram illustrating patient attrition.**

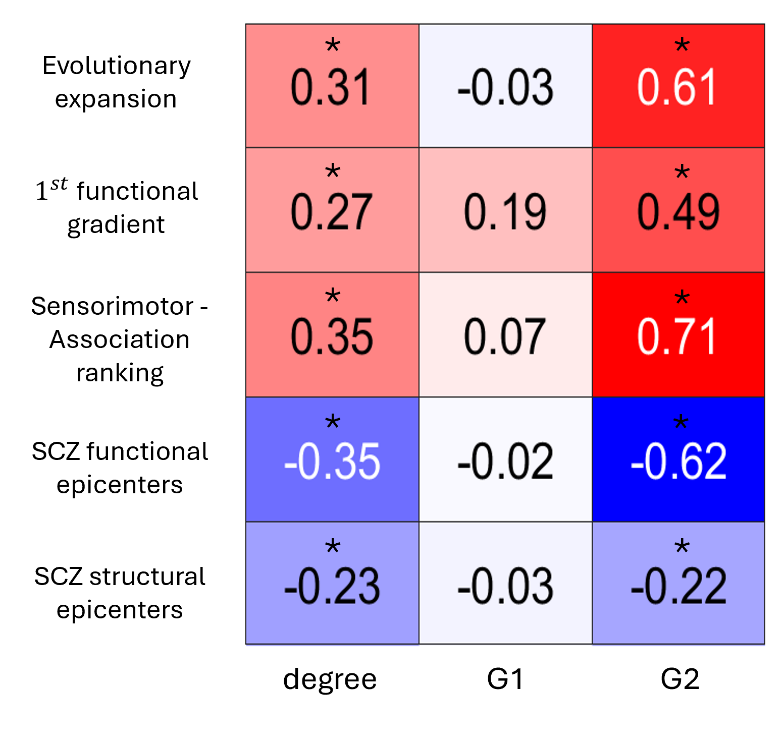

**Supplementary Fig. 3 Regional co-localization of MIND (degree, G1, and G2) in SSD with brain maps of cortical hierarchy and SCZ epicenters.** Asterisks (*) indicate significant correlations (Benjamini-Hochberg false discovery rate (FDR)-corrected across features; *P*_spin_ < 0.05). G1, first gradient; G2, second gradient; SCZ, schizophrenia.

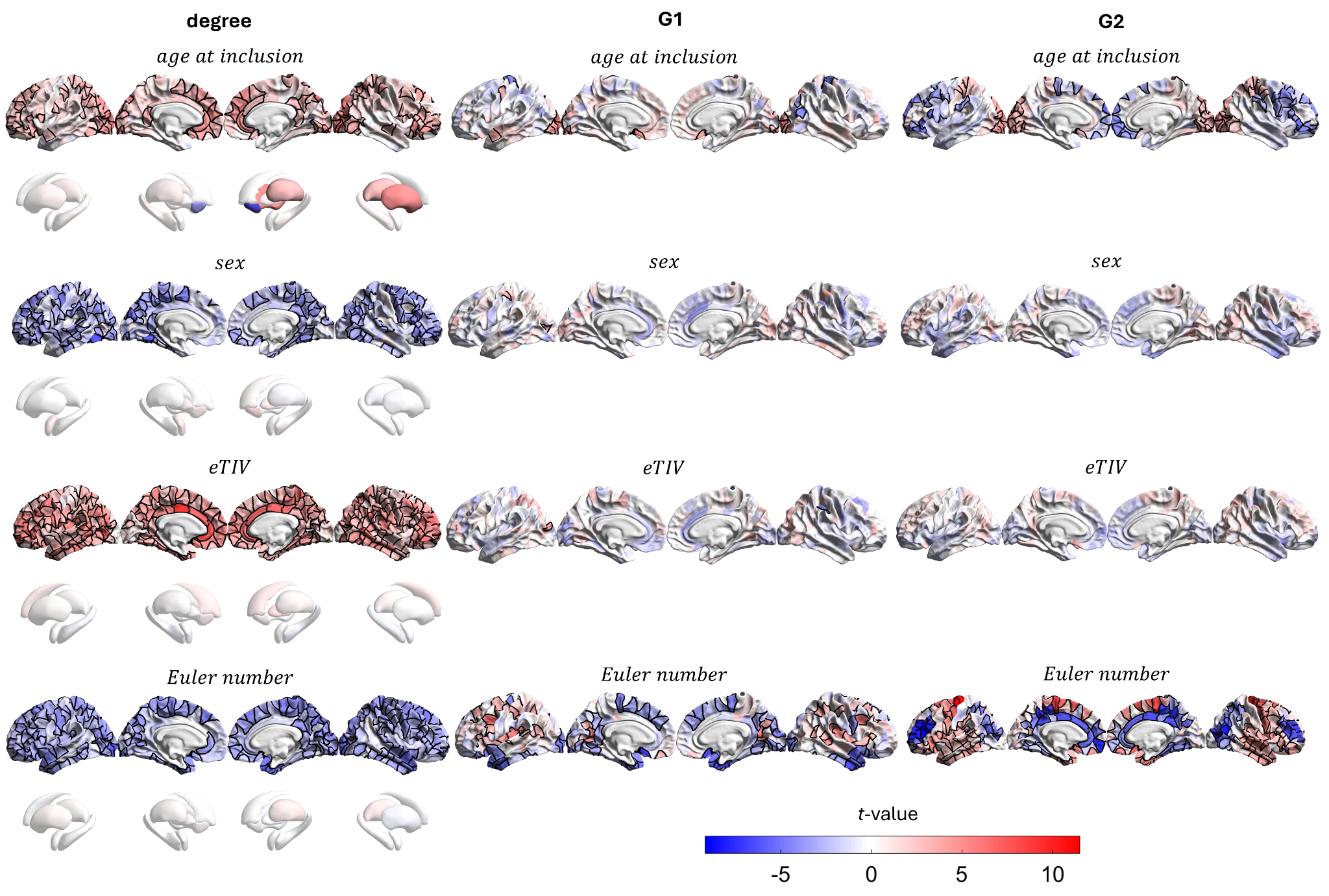

**Supplementary Fig. 4 Baseline effects of age at inclusion, sex, eTIV, and Euler number on MIND degree and gradients in SSD.** Regional associations (*t*-values) representing the contribution of each variable on brain similarity assessed at baseline by multiple regression models (from left to right: MIND, G1, and G2; FDR-corrected, *P* < 0.05). eTIV, estimated total intracranial volume; G1, first gradient; G2, second gradient.

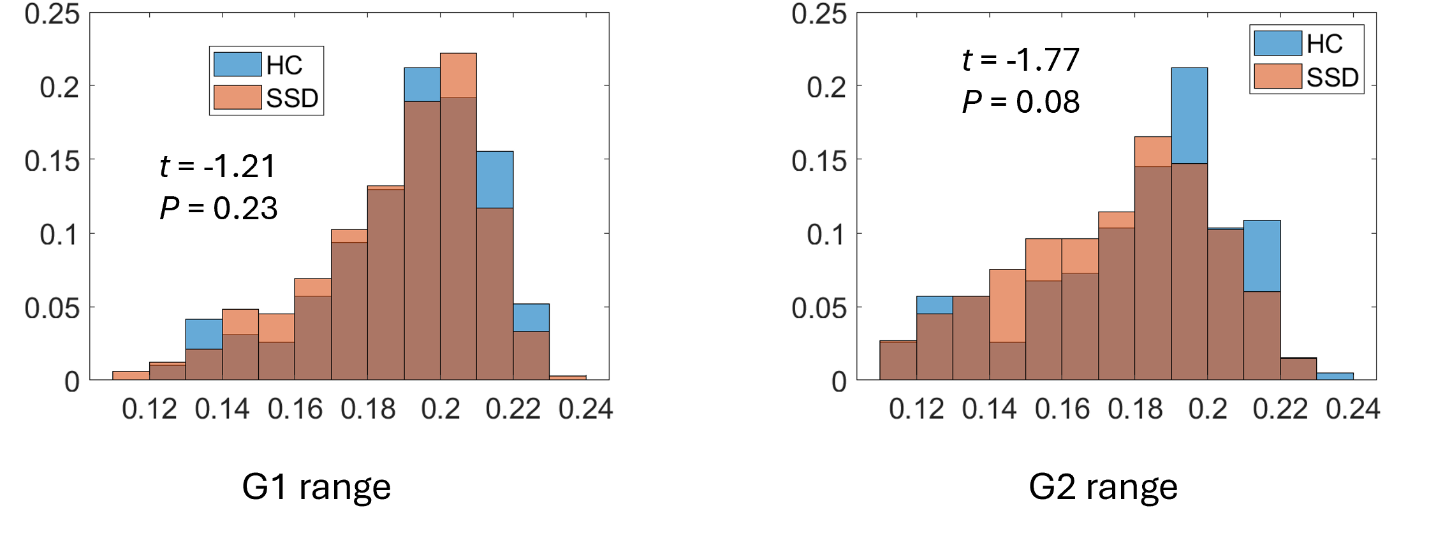

**Supplementary Fig. 5 Histograms of G1 and G2 gradient ranges.** Baseline effects (*t*-values) of SSD diagnosis on G1 and G2 gradient ranges assessed by multiple regression models after accounting for age at inclusion, sex, eTIV, Euler number. eTIV, estimated total intracranial volume; G1, first gradient; G2, second gradient.

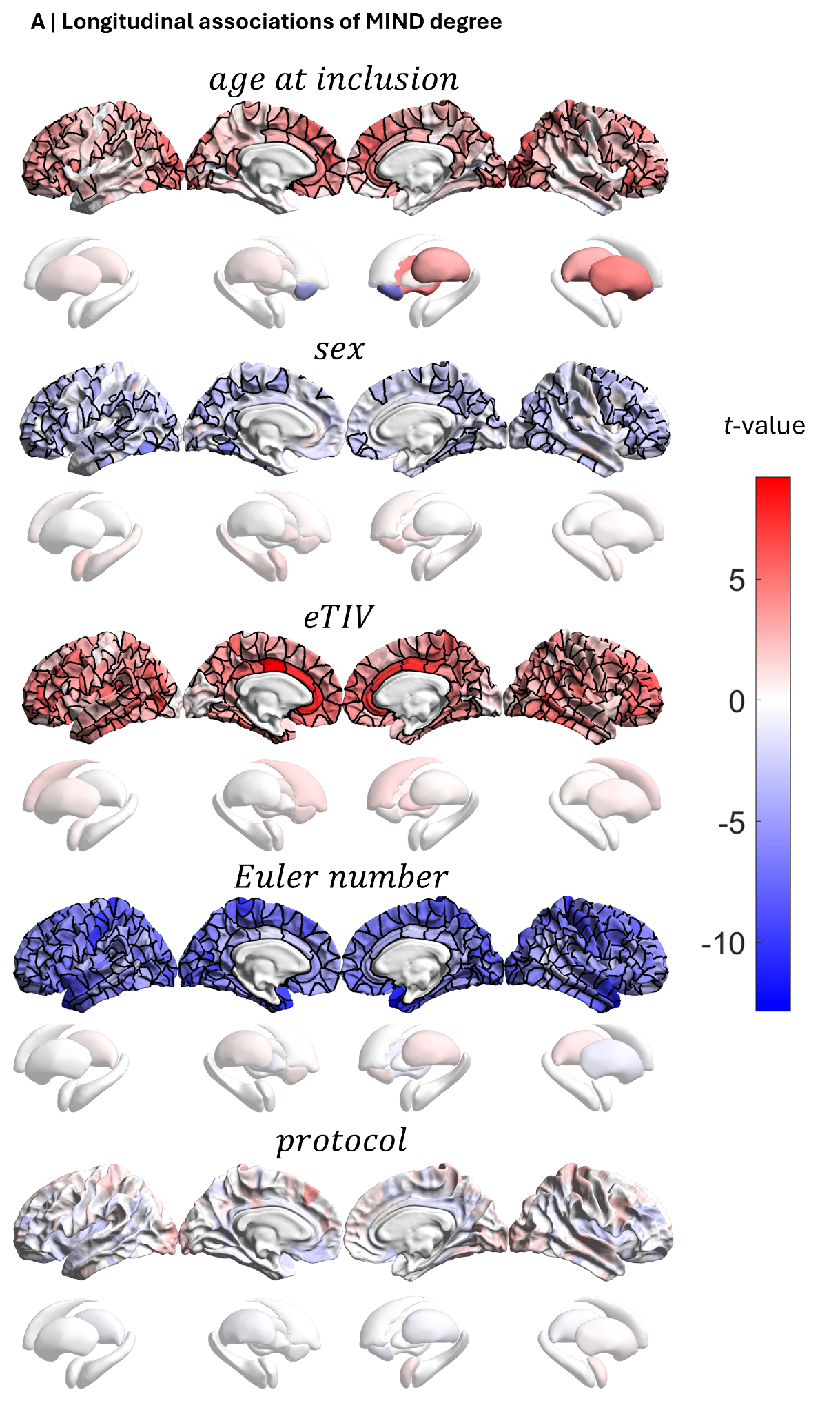

**Supplementary Fig. 6 Longitudinal effects of age at inclusion, sex, eTIV, Euler number, and protocol on MIND degree in SSD.** Regional *t*-values representing the contribution of each variable to the linear mixed model (LMM) assessing the longitudinal relationship between MIND networks and covariates in SSD (FDR-corrected, *P* < 0.05). eTIV, estimated total intracranial volume; MIND, Morphometric INverse Divergence.

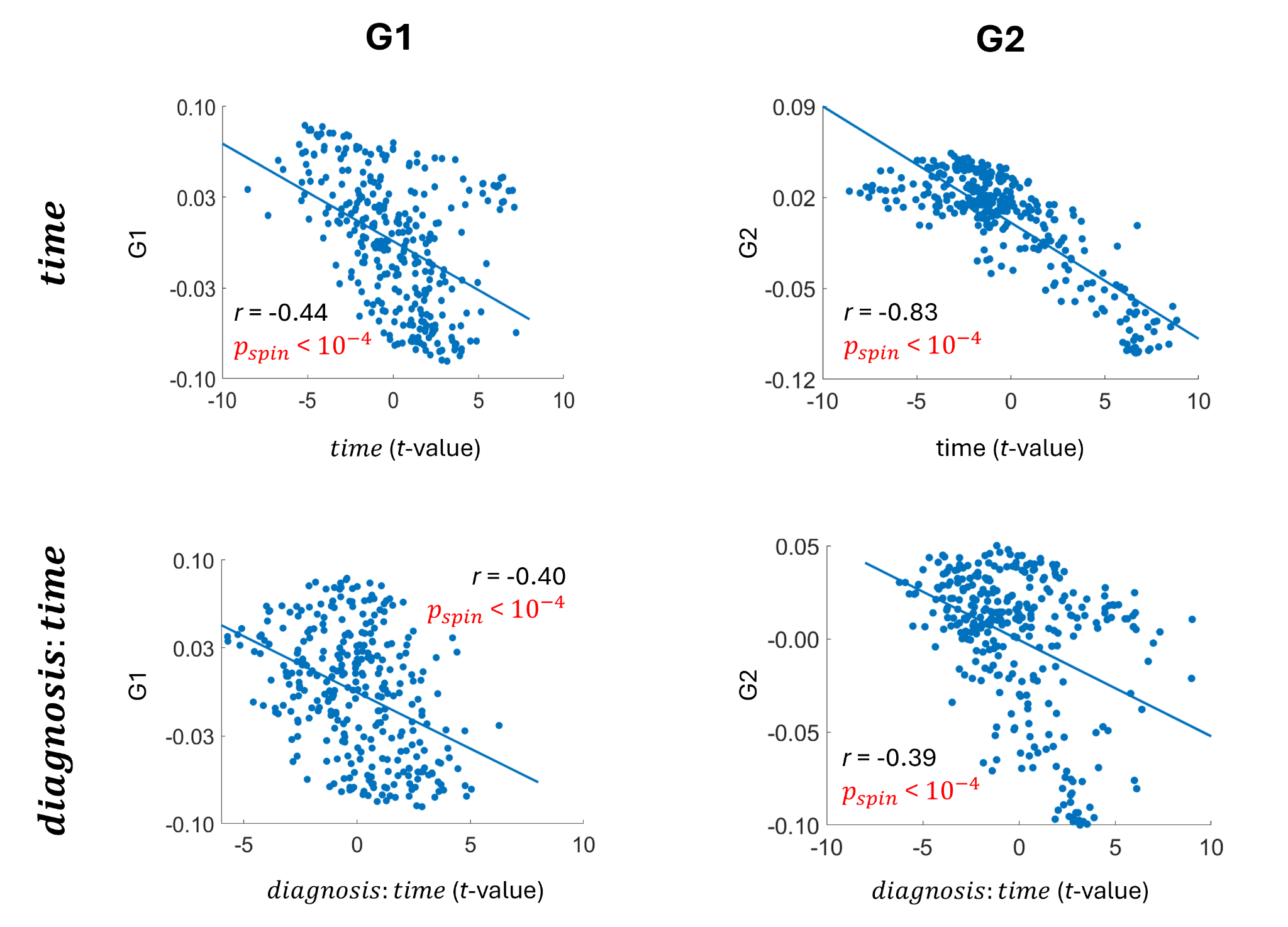

**Supplementary Fig. 7 Scatter plots representing gradient compression (a reduction in regional differentiation) across time.** Regional correlations (two-sided test) between HC gradients and the effect of *time* and *diagnosis:time* on gradients. G1, first gradient; G2, second gradient.

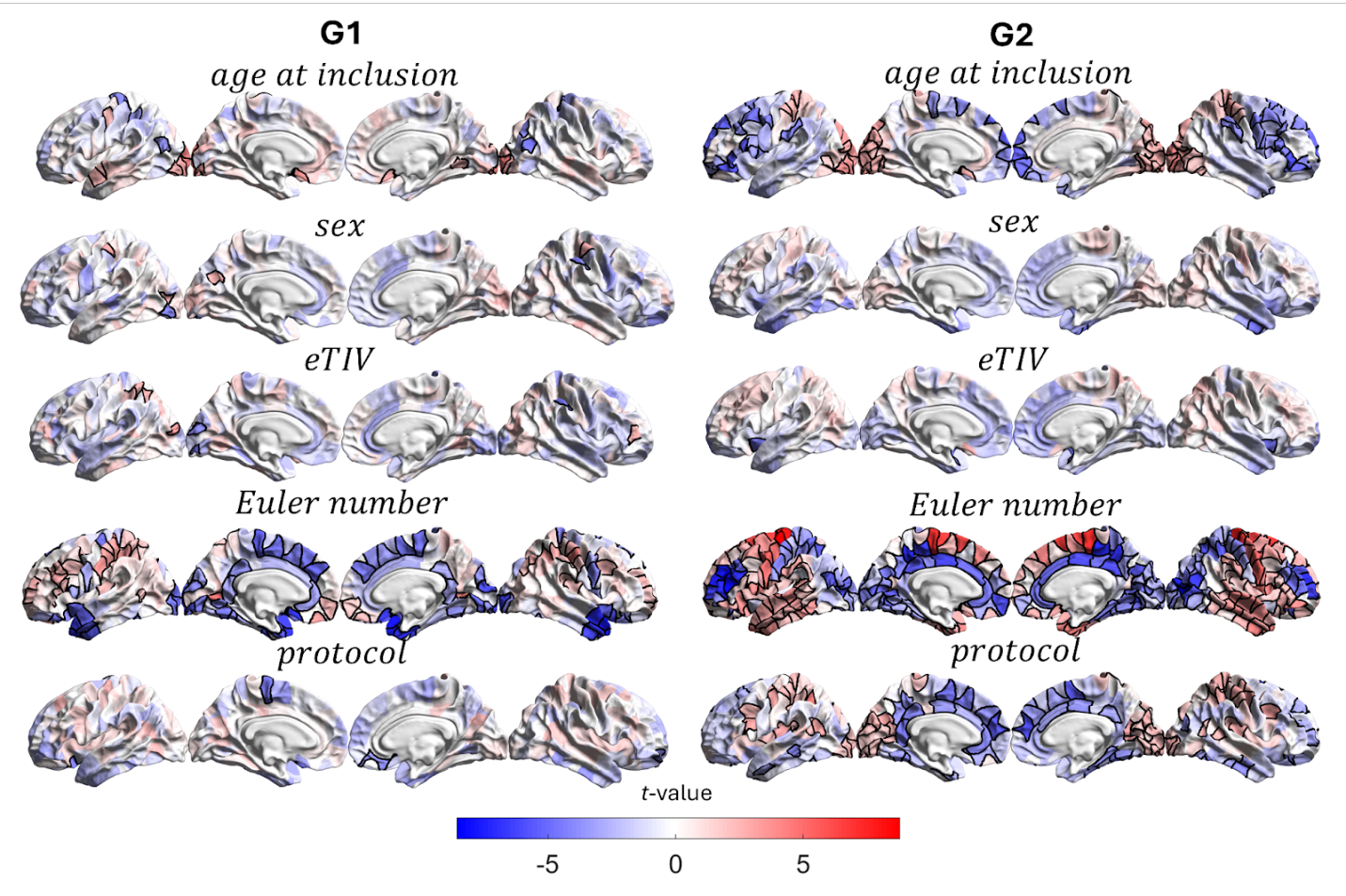

**Supplementary Fig. 8 Longitudinal effects of age at inclusion, sex, eTIV, Euler number, and protocol on MIND gradients in SSD.** Regional *t*-values representing the contribution of each variable to the LMM assessing the longitudinal relationship between structural gradients and covariates in SSD (FDR-corrected, *P* < 0.05). eTIV, estimated total intracranial volume; G1, first gradient; G2, second gradient.

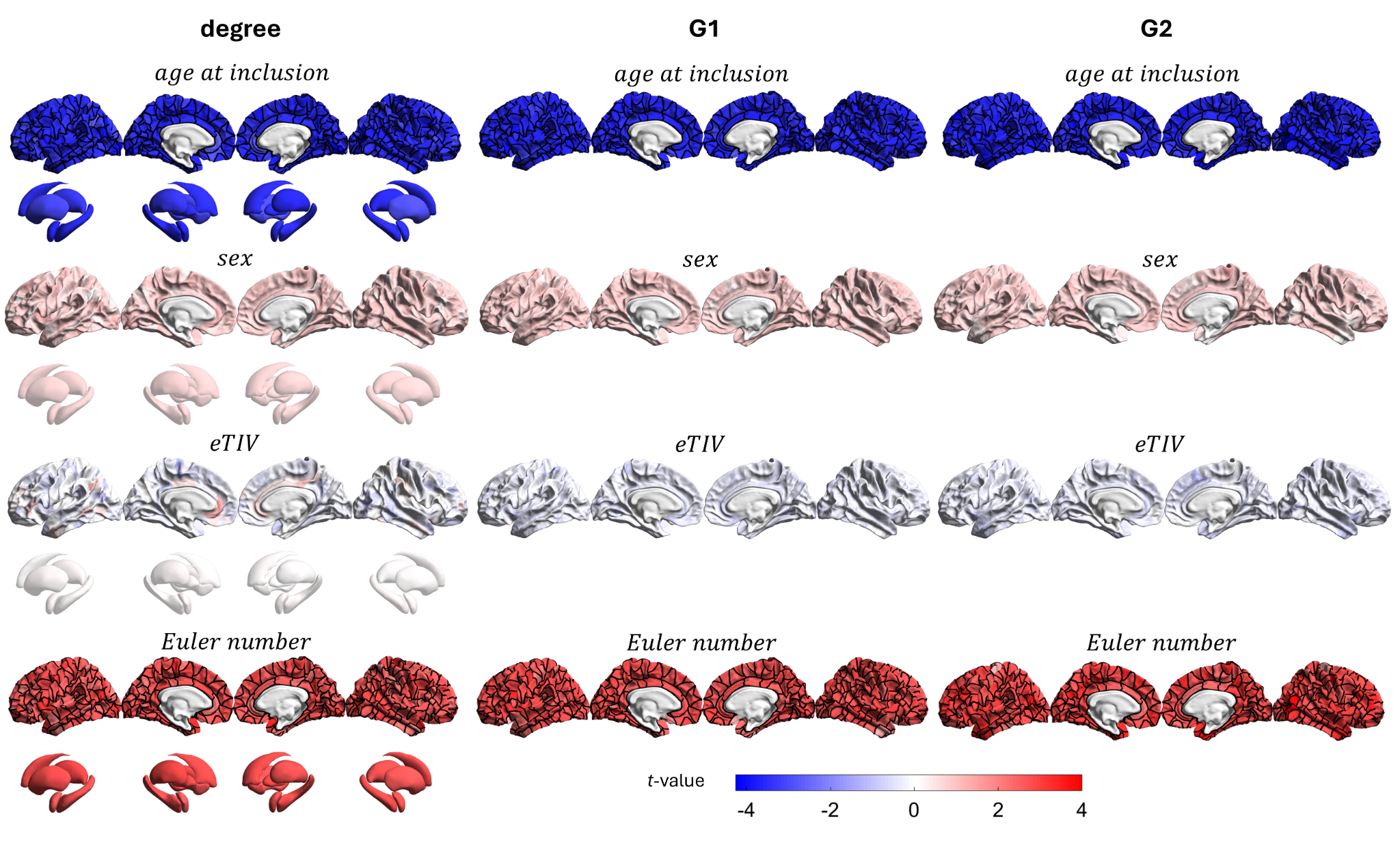

**Supplementary Fig. 9 Baseline effects of age at inclusion, sex, eTIV, and Euler number on psychiatric symptoms in SSD.** Regional *t*-values representing the contribution of each variable to the GLM assessing the baseline relationship between psychiatric symptoms and covariates (FDR-corrected, *P* < 0.05). eTIV, estimated total intracranial volume; G1, first gradient; G2, second gradient.

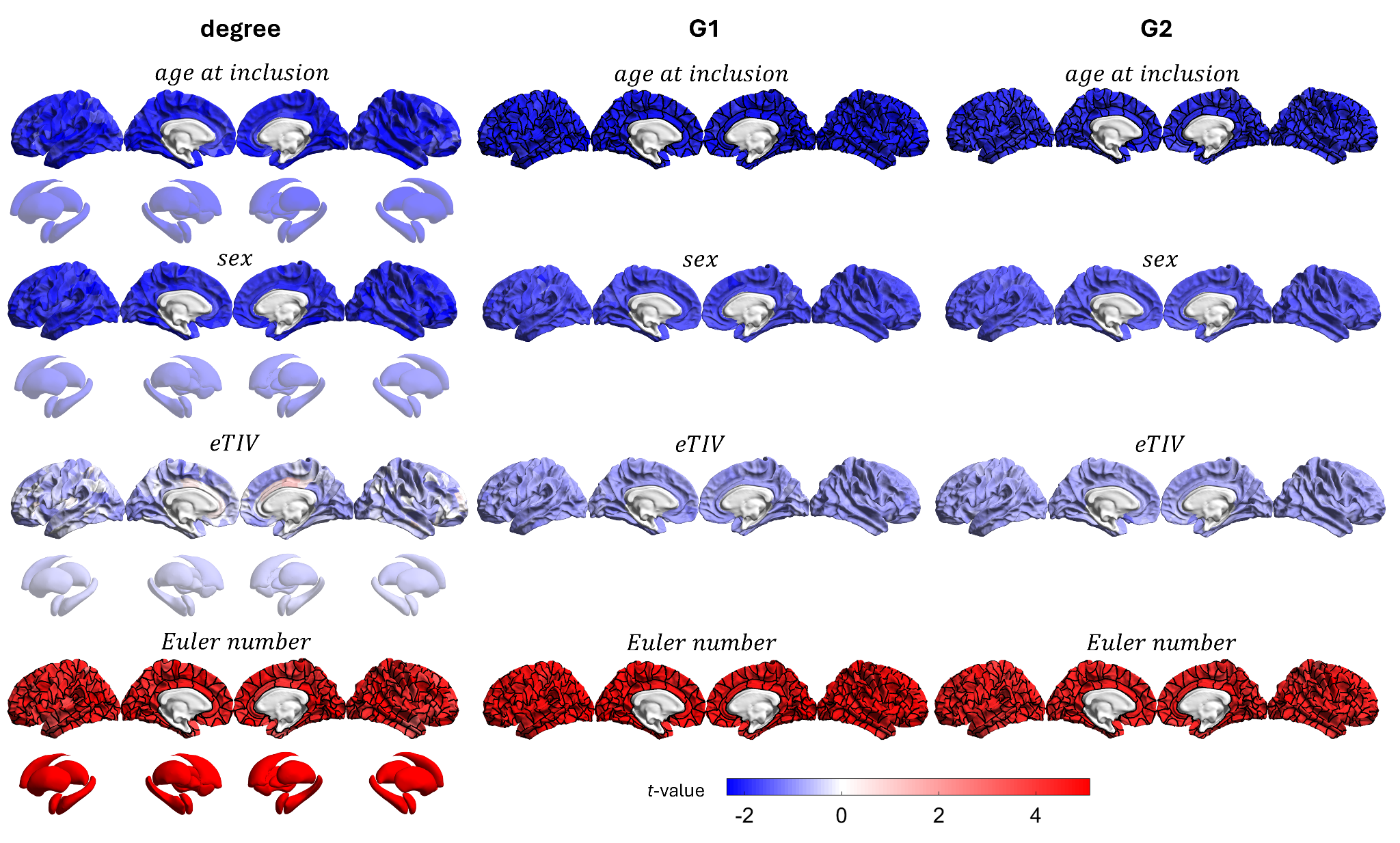

**Supplementary Fig. 10 Longitudinal effects of age at inclusion, sex, eTIV, and Euler number on psychiatric symptoms in SSD.** Regional *t*-values representing the contribution of each variable to the GLMM assessing the longitudinal relationship between psychiatric symptoms and covariates (FDR-corrected, *P* < 0.05). eTIV, estimated total intracranial volume; G1, first gradient; G2, second gradient.

### Supplemental References

1.	Crespo-Facorro B, Pérez-Iglesias R, Ramirez-Bonilla M, Martínez-García O, LLorca J, Vázquez-Barquero JL. A Practical Clinical Trial Comparing Haloperidol, Risperidone, and Olanzapine for the Acute Treatment of First-Episode Nonaffective Psychosis. J Clin Psychiatry. 2006;67:1511–1521.

2.	Rodriguez-Perez N, Ayesa-Arriola R, Ortiz-García de la Foz V, Setien-Suero E, Tordesillas-Gutierrez D, Crespo-Facorro B. Long term cortical thickness changes after a first episode of non- affective psychosis: The 10 year follow-up of the PAFIP cohort. Progress in Neuro-Psychopharmacology and Biological Psychiatry. 2021;108:110180.

3.	Andreasen NC, Flaum M, Arndt S. The Comprehensive Assessment of Symptoms and History (CASH): An Instrument for Assessing Diagnosis and Psychopathology. Archives of General Psychiatry. 1992;49:615–623.

4.	Vos de Wael R, Benkarim O, Paquola C, Lariviere S, Royer J, Tavakol S, et al. BrainSpace: a toolbox for the analysis of macroscale gradients in neuroimaging and connectomics datasets. Commun Biol. 2020;3:103.

5.	Luppi AI, Uhrig L, Tasserie J, Signorelli CM, Stamatakis EA, Destexhe A, et al. Local orchestration of distributed functional patterns supporting loss and restoration of consciousness in the primate brain. Nat Commun. 2024;15:2171.

6.	Langs G, Golland P, Ghosh SS. Predicting Activation Across Individuals with Resting-State Functional Connectivity Based Multi-Atlas Label Fusion. Med Image Comput Comput Assist Interv. 2015;9350:313–320.

7.	Hong S-J, Vos de Wael R, Bethlehem RAI, Lariviere S, Paquola C, Valk SL, et al. Atypical functional connectome hierarchy in autism. Nat Commun. 2019;10:1022.

8.	He Y, Li Q, Fu Z, Zeng D, Han Y, Li S. Functional gradients reveal altered functional segregation in patients with amnestic mild cognitive impairment and Alzheimer’s disease. Cereb Cortex. 2023;33:10836–10847.

9.	Inada T, Inagaki A. Psychotropic dose equivalence in Japan. Psychiatry and Clinical Neurosciences. 2015;69:440–447.

10.	Gardner DM, Murphy AL, O’Donnell H, Centorrino F, Baldessarini RJ. International consensus study of antipsychotic dosing. Am J Psychiatry. 2010;167:686–693.

11.	Hill J, Inder T, Neil J, Dierker D, Harwell J, Van Essen D. Similar patterns of cortical expansion during human development and evolution. Proc Natl Acad Sci U S A. 2010;107:13135–13140.

12.	Margulies DS, Ghosh SS, Goulas A, Falkiewicz M, Huntenburg JM, Langs G, et al. Situating the default-mode network along a principal gradient of macroscale cortical organization. Proceedings of the National Academy of Sciences. 2016;113:12574–12579.

13.	Sydnor VJ, Larsen B, Bassett DS, Alexander-Bloch A, Fair DA, Liston C, et al. Neurodevelopment of the association cortices: Patterns, mechanisms, and implications for psychopathology. Neuron. 2021;109:2820–2846.

14.	Georgiadis F, Larivière S, Glahn D, Hong LE, Kochunov P, Mowry B, et al. Connectome architecture shapes large-scale cortical alterations in schizophrenia: a worldwide ENIGMA study. Mol Psychiatry. 2024;29:1869–1881.

15.	Markello RD, Misic B. Comparing spatial null models for brain maps. NeuroImage. 2021;236:118052.

16.	Váša F, Mišić B. Null models in network neuroscience. Nat Rev Neurosci. 2022;23:493–504.
